# Income, access to care and adult oral health inequalities in the United States: a multilevel analysis of national surveys and Medicaid policies

**DOI:** 10.64898/2025.12.12.25342177

**Authors:** Zhou Chengke

## Abstract

**Background:** Socioeconomic status (SES) strongly shapes oral health, yet the magnitude of these gradients, their pathways and the influence of US Medicaid policies remain uncertain. We quantified SES gradients in adult oral health, examined potential mechanisms and assessed whether state Medicaid expansion and adult dental benefit generosity were associated with aggregate oral health indicators.

**Methods:** We analysed adults aged ≥20 years in NHANES 1999–2019 and state adult populations in BRFSS 2011–2025, supplemented with international oral health and Medicaid policy data. Individual outcomes were DMFT and self-rated oral health (good vs fair/poor). State-level outcomes were past-year dental visit rates, any permanent tooth loss and complete edentulism among adults ≥65 years. SES measures included poverty–income ratio (PIR) and education; mediators included annual dental visits, unmet dental need and sleep duration. Analytic methods comprised survey-weighted regression, concentration and slope indices of inequality, Oaxaca–Blinder decomposition and state-level difference-in-differences models with state and year fixed effects and state-clustered standard errors.

**Results:** Higher PIR and education were independently associated with lower DMFT and higher odds of good self-rated oral health in all age groups. PIR coefficients for DMFT were ≈−0.25 (ages 20–44), −0.79 (45–64) and −1.07 (≥ 65); corresponding odds ratios for good oral health were ≈1.48, 1.46 and 1.33. Predicted probabilities of good oral health increased monotonically across PIR quartiles. Concentration indices indicated that DMFT burden was concentrated among low-income adults (CI ≈ −0.105), whereas good oral health was concentrated among high-income adults (CI ≈ 0.094). The Slope Index suggested that moving from the lowest to highest income rank corresponded to ≈2.48 fewer affected teeth; the Relative Index indicated ≈eight-fold higher odds of reporting good oral health. Oaxaca–Blinder decomposition showed a Q4–Q1 DMFT gap of 1.31 teeth, with roughly one quarter explained by observed variables, mainly differences in dental access. State-level difference-in-differences models did not identify large, precisely estimated changes in dental visit rates, tooth loss or edentulism associated with Medicaid expansion or adult dental benefit generosity.

**Conclusion:** Marked SES-related oral health inequalities persist among US adults, particularly in midlife, and are strongly linked to differential dental access and socially patterned behaviours. Medicaid expansion and adult dental benefit generosity, as implemented, did not produce substantial detectable shifts in state-level oral health indicators. Reducing inequalities will require improved financial protection for dental care and broader action on income, education and other social determinants of health.

## 1 Introduction

### 1.1 Global and national context of oral health inequalities

Oral health is a fundamental component of general health, well-being and social participation[1]. Untreated dental caries, severe periodontal disease and edentulism are among the most prevalent non-communicable conditions worldwide and can impair chewing, speech, appearance and quality of life. Despite advances in preventive technologies and restorative care, the global distribution of oral health and oral health services remains highly unequal.

High-income countries generally have more dentists per capita, greater use of preventive care and lower levels of untreated caries than many low- and middle-income countries, yet pronounced social gradients persist within almost all countries[1, 2].

China and the United States illustrate this tension between overall improvement and persistent inequality. In China, successive national oral health surveys show that, alongside rising awareness and some gains in prevention, large urban–rural, regional and socioeconomic disparities remain in caries, periodontal disease and tooth loss[3, 4]. In the United States, analyses of the National Health and Nutrition Examination Survey (NHANES) consistently find that adults with lower income and education have higher DMFT scores, worse self-rated oral health and greater functional limitation. In both countries, adult dental services are only partly covered by public schemes, and many households face substantial out-of-pocket spending. Macro-level WHO data suggest that higher national income and health expenditure are associated with better average oral health, but these relationships are non-linear and sensitive to age structure and data quality, underscoring the need for individual-level analyses that clarify how socioeconomic status and health system design combine to shape oral health inequalities[3, 5].

### 1.2 Conceptual framework: pathways from socioeconomic status to oral health

The social determinants of health framework provides a useful lens for understanding how socioeconomic status (SES) translates into oral health inequalities. At least four interrelated pathways can be distinguished[6].

Socioeconomic status (SES) influences oral health through four overlapping pathways[7–9]. Material conditions determine whether people can afford transport, hygiene products and dental treatment. Behavioural and cultural factors, shaped by education and social position, affect toothbrushing, diet, smoking, sleep and dental visiting. Psychosocial stress and insecurity can dysregulate physiology and promote harmful coping behaviours[10, 11].

Institutional and service arrangements, including insurance coverage and provider distribution, structure access to care. These pathways interact over the life course and are shaped by geography[12], race/ethnicity and policy decisions about whether dental care is included in basic health coverage.1.3 Study objectives

Existing studies have described socioeconomic gradients in oral health in many countries, but important gaps remain[5, 13, 14]. First, relatively few analyses quantify the magnitude of income-related inequality using summary indices such as the concentration index (CI), Slope Index of Inequality (SII) and Relative Index of Inequality (RII), or compare these across age groups. Second, although access to dental care and health behaviours are widely recognised as important mediators, there is limited work that formally decomposes their contribution to socioeconomic gaps in cumulative oral disease. Third, while there is a growing literature on the effects of Affordable Care Act (ACA) Medicaid expansion and changes in adult Medicaid dental benefits, their implications for population-level oral health indicators remain uncertain. Finally, evidence from the United States is rarely interpreted alongside findings from China and other countries seeking to integrate oral health into universal health coverage[8, 15].

Against this background, the present study aims to provide a comprehensive assessment of socioeconomic inequalities in oral health among US adults and to situate these findings in a broader international context. Specifically, we:

1. describe and quantify gradients in both objective (DMFT) and subjective (self-rated oral health) outcomes across income and education levels, overall and by age group, using nationally representative NHANES data;
2. compute CI, SII and RII to summarise income-related inequalities in oral health and compare their magnitude across adult age groups;
3. examine potential mechanisms by conducting mediation analysis and Oaxaca–Blinder decomposition to assess the contribution of dental care access (annual visits and unmet need) and sleep-related behaviours to socioeconomic gaps in oral health[16–18]; and
4. construct a state–year panel of Medicaid expansion status and adult Medicaid dental benefit generosity, link it to BRFSS state-level indicators of dental visits and tooth loss, and use difference-in-differences models to explore whether recent Medicaid policy changes are associated with shifts in aggregate oral health indicators.

Macro-level WHO data on oral health and health system indicators are used descriptively in the Introduction and Discussion to illustrate cross-national patterns and to contrast the complex relationships seen at the country level with the clearer gradients observed in individual-level analyses.

## 2 Methods

### 2.1 Data sources

We combined three main data sources to examine socioeconomic inequalities in oral health in the United States and their policy context. First, we used individual-level data from the National Health and Nutrition Examination Survey (NHANES), a continuous, nationally representative survey of the US civilian, non-institutionalised population that includes standardised interviews and clinical oral examinations. Second, we used state-level data from the Behavioral Risk Factor Surveillance System (BRFSS), a state-based telephone survey that provides age-adjusted prevalence estimates for health behaviours and outcomes, including oral health indicators, for each US state and year. Third, we constructed a state–year panel of Medicaid policies using publicly available documentation from the Kaiser Family Foundation (KFF), the Centre for Health Care Strategies (CHCS), the Medicaid and CHIP Payment and Access Commission (MACPAC) and CareQuest Institute reports, which summarise Affordable Care Act (ACA) Medicaid expansion status and adult Medicaid dental benefit generosity by state and year.

To provide international context, we also drew on country-level data from World Health Organization (WHO) oral health databases estimates. These macro-level data were used descriptively in the Introduction and Discussion to illustrate cross-national patterns in oral disease burden, dental workforce capacity and health expenditure. Detailed variable definitions and sensitivity analyses based on WHO are presented in the Supplementary materials.

### 2.2 Measures

#### 2.2.1 NHANES outcomes, exposures and covariates

The primary objective outcome in NHANES was the decayed, missing and filled teeth index (DMFT), derived from clinical dental examinations and treated as a continuous count of affected teeth. The main subjective outcome was self-rated oral health, assessed on a five-point scale (excellent, very good, good, fair, poor) and dichotomised as good/very good/excellent versus fair/poor.

Socioeconomic status (SES) was measured using the poverty-income ratio (PIR) and educational attainment. PIR is defined as the ratio of family income to the federal poverty threshold for the corresponding year and family size; we analysed PIR both as a continuous variable and in quartiles (Q1 lowest to Q4 highest). Education was categorised as less than high school, high school or General Educational Development (GED), some college or associate degree, and college degree or higher.

We examined two sets of potential mediators. First, access to dental care was captured by: (1) having visited a dentist or dental clinic in the past 12 months for any reason (yes/no); and (2) reporting unmet dental need in the past year (yes/no), defined as needing dental care but not obtaining it. Among respondents with unmet dental need, we further examined self-reported main reasons, focusing on cost and being “too busy / no time” as key domains.

Second, behavioural and sleep-related factors included short sleep duration (e.g. <6 hours per night) and selected behaviours such as smoking status that are plausibly associated with oral health and available consistently across survey cycles. Additional behaviours (e.g. consumption of sugar-sweetened beverages[19, 20], toothbrushing frequency) were incorporated in exploratory principal component analyses and sensitivity analyses described in the Supplement.

Covariates included age, sex, race/ethnicity and other demographic characteristics. Age was modelled both continuously (per 10-year increase) and categorically in three groups: 20–44, 45–64 and ≥65 years. Race/ethnicity was grouped into non-Hispanic White, non-Hispanic Black, Hispanic and other racial/ethnic groups according to NHANES definitions. Additional covariates such as marital status, nativity and general health status were considered in sensitivity analyses. Unless otherwise specified, analyses were restricted to participants aged ≥20 years with complete data on outcomes, PIR, education, dental access variables and key covariates.

#### 2.2.2 BRFSS outcomes and Medicaid policy variables

From BRFSS we derived three age-adjusted, state-level oral health indicators for each calendar year: (1) the proportion of adults reporting a dental visit in the past 12 months for any reason (dental_visit_rate); (2) the proportion of adults reporting loss of at least one permanent tooth due to decay or periodontal disease (any_tooth_loss_rate); and (3) the proportion of adults aged ≥65 years reporting complete loss of all natural teeth (edentulism_65_rate).

Medicaid policy variables were constructed from KFF, CHCS, MACPAC and CareQuest sources. For each state-year, we created an indicator of ACA Medicaid expansion status (MedicaidExpansion_st), coded as 1 in years in which coverage under the ACA Medicaid expansion was in effect and 0 otherwise. Adult Medicaid dental benefit generosity (DentalGenerosityLevel_st) was coded as an ordinal variable taking values 0 (no or emergency-only dental coverage), 1 (limited coverage) and 2 (extensive coverage), based on harmonised classifications of benefit scope and coverage level. When sources disagreed or benefit changes occurred mid-year, we followed the most recent comprehensive synthesis and coded the year according to the predominant policy in effect.

### 2.3 Statistical analysis

#### 2.3.1 Descriptive analyses

All NHANES analyses incorporated examination weights, strata and primary sampling units to account for the complex, multistage sampling design. We first described weighted distributions of DMFT, self-rated oral health, dental visits, unmet dental need and reasons for unmet need by PIR quartile and education level, overall and within age groups (20–44, 45–64 and ≥65 years). For BRFSS, we examined temporal trends in state-level dental visit rates, any tooth loss and edentulism among older adults, stratified by Medicaid expansion status and adult dental benefit generosity categories.

#### 2.3.2 NHANES regression models

We estimated survey-weighted linear regression models with DMFT as the dependent variable and survey-weighted logistic regression models with self-rated good oral health as the dependent variable. Main exposure variables were PIR (continuous) and education categories. Models were adjusted for age, sex, race/ethnicity and other covariates. To assess how SES gradients varied across the adult life course, we fitted age-stratified models for adults aged 20–44, 45–64 and ≥65 years. From the logistic models we derived model-based predicted probabilities of self-rated good oral health for each PIR quartile within age groups.

#### 2.3.3 Income-related inequality indices

To summarise income-related inequalities in oral health, we calculated three complementary indices. The concentration index (CI) was computed for DMFT (a “bad” outcome) and self-rated good oral health (a “good” outcome), using the fractional rank of PIR in the income distribution. Negative CI values for DMFT indicate that higher disease burden is concentrated among poorer individuals, whereas positive CI values for self-rated good oral health indicate concentration among richer individuals.

The Slope Index of Inequality (SII) for DMFT was estimated by regressing DMFT on the relative rank of PIR (from 0 for the lowest to 1 for the highest) with adjustment for age, sex and race/ethnicity. The SII represents the absolute difference in DMFT between the top and bottom of the income hierarchy. The Relative Index of Inequality (RII) for self-rated good oral health was estimated using logistic regression models of the log-odds of reporting good oral health on PIR rank, providing an odds ratio comparing the highest versus lowest position in the income distribution. We computed CI, SII and RII for the overall adult population and separately for the three age groups[21].

#### 2.3.4 Mediation analysis and Oaxaca–Blinder decomposition

To explore potential mechanisms linking SES to oral health, we conducted mediation analyses with PIR or education as the exposure, DMFT or self-rated good oral health as the outcome, and dental access variables (annual dental visit and unmet dental need) and short sleep duration as candidate mediators[17, 22]. Indirect and direct effects were estimated using product-of-coefficients approaches within survey-weighted regression frameworks. We used non-parametric bootstrap resampling to obtain 95% confidence intervals; intervals that did not cross zero were interpreted as evidence of statistically significant mediation.

We further applied a twofold Oaxaca–Blinder decomposition to differences in mean DMFT between adults in the lowest (Q1) and highest (Q4) PIR quartiles. Survey-weighted linear regression models for DMFT were fitted separately within Q1 and Q4. Explanatory variables were grouped into domains: (1) access to dental care (annual dental visit and unmet dental need); (2) behavioural and sleep-related factors; (3) education; (4) demographic characteristics (age in decades and sex); and (5) race/ethnicity. The total Q4–Q1 difference in mean DMFT was decomposed into an “explained” component attributable to differences in observed characteristics across PIR quartiles and an “unexplained” component reflecting differences in coefficients and unmeasured factors. Domain-level contributions were obtained by summing contributions across variables within each domain.

#### 2.3.5 State-level difference-in-differences models

To assess whether Medicaid expansion and adult Medicaid dental benefit generosity were associated with changes in state-level oral health indicators, we estimated difference-in-differences (DID) models of the form:

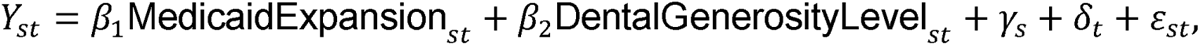

where *Y_st_* denotes each BRFSS outcome in state and year *t*; *γ_s_* are state fixed effects capturing time-invariant differences across states; *δ_t_* are year fixed effects capturing common national trends; and *ε_st_* is the error term. The coefficients *β*_1_ and *β*_2_ represent average changes in the outcome associated with Medicaid expansion and with higher levels of adult Medicaid dental benefit generosity, respectively, under standard DID assumptions. Models were estimated using ordinary least squares with standard errors clustered at the state level. Our primary DID outcome was dental_visit_rate as a proximal indicator of access to care; any_tooth_loss_rate and edentulism_65_rate were examined as more distal, exploratory outcomes..

All statistical analyses were conducted using standard software (R, Python and Stata). Two-sided p values less than 0.05 were considered statistically significant, and 95% confidence intervals are reported where appropriate.

## 3 Results

### 3.1 Sample characteristics and access to dental care

The pooled NHANES sample included adults aged 20 years and older across multiple survey cycles. Socioeconomically disadvantaged participants tended to be younger, more likely to be non-White, and to have lower dental visit rates, higher unmet dental need and worse oral health indicators than their more advantaged counterparts (Table 1).

**Table 1.** Weighted characteristics of adult NHANES participants by poverty-income ratio (PIR) quartile, 1999–2019. Values are survey-weighted means (standard errors) or percentages unless otherwise specified. PIR, poverty-income ratio; DMFT, decayed, missing and filled teeth.

| Characteristic | Overall | PIR Q1<br>(lowest) | PIR Q2 | PIR Q3 | PIR Q4<br>(highest) |
| --- | --- | --- | --- | --- | --- |
| Unweighted N | 60207 | 20280 | 16768 | 12509 | 10650 |
| Weighted % female | 51.9 | 56.4 | 52.7 | 50.5 | 48.0 |
| Mean age (years) | 47.1 | 44.5 | 48.2 | 46.9 | 48.6 |
| Age 20-44 (%) | 47.4 | 55.5 | 47.1 | 48.1 | 38.9 |
| Age 45-64 (%) | 34.3 | 27.1 | 29.1 | 34.4 | 46.8 |
| Age ≥65 (%) | 18.3 | 17.4 | 23.7 | 17.6 | 14.4 |
| NH White (%) | 67.1 | 50.6 | 62.1 | 73.7 | 82.3 |
| NH Black (%) | 11.3 | 17.0 | 13.2 | 9.1 | 5.8 |
| Hispanic (%) | 14.1 | 24.4 | 16.8 | 10.1 | 4.9 |
| Other<br>race/ethnicity (%) | 7.5 | 8.1 | 7.9 | 7.1 | 7.0 |
| Education <HS (%) | 10.1 | 22.3 | 12.6 | 4.4 | 1.9 |
| Education HS/GED (%) | 26.5 | 39.4 | 32.3 | 23.8 | 10.6 |
| Education some | 29.4 | 28.3 | 31.6 | 35.9 | 21.5 |
| college (%) |  |  |  |  |  |
| Education >=college (%) | 34.0 | 10.0 | 23.6 | 36.0 | 66.0 |
| Mean PIR | 2.90 | 0.92 | 2.02 | 3.66 | 4.99 |
| Mean DMFT (SE) | 3.94<br>(0.033) | 4.93<br>(0.067) | 4.36<br>(0.068) | 3.62<br>(0.063) | 2.85<br>(0.048) |
| % self-rated good oral health | 72.2 | 55.8 | 68.0 | 77.9 | 87.2 |
| % past-year dental visit | 74.4 | 62.0 | 71.3 | 77.1 | 87.5 |
| % unmet dental need | 8.7 | 16.3 | 9.6 | 5.9 | 3.0 |

Across survey cycles, approximately two thirds of adults reported at least one dental visit in the past 12 months, but there was a clear gradient by income. Past-year dental visiting was substantially less common in the lowest poverty-income ratio (PIR) quartile than in the highest, with absolute differences of around 20–30 percentage points. Unmet dental need was also strongly patterned by SES. Overall, about 14–16% of adults reported that they needed dental care but did not obtain it in the previous year, but this proportion reached roughly 25–30% among adults in the lowest PIR quartile compared with about 5–10% in the highest quartile. Similar patterns were observed when stratifying by education level, with adults who had not completed high school exhibiting the highest levels of unmet need.

### 3.2 Socioeconomic gradients in DMFT and self-rated oral health

In survey-weighted regression models, higher income and higher education were consistently associated with better oral health. After adjustment for age, sex, race/ethnicity and other covariates, PIR showed a negative association with DMFT and a positive association with self-rated good oral health in the overall adult sample and within all age strata (Table 2). In age-stratified linear models, the estimated PIR coefficients for DMFT were approximately −0.25 for adults aged 20–44 years, −0.79 for those aged 45–64 years and −1.07 for adults aged ≥65 years, indicating progressively larger absolute reductions in cumulative caries and tooth loss with higher income at older ages. In corresponding logistic models, the odds ratios for self-rated good oral health per unit increase in PIR were around 1.48, 1.46 and 1.33 in the three age groups, respectively. Higher education levels were independently associated with lower DMFT and higher odds of reporting good oral health, even after controlling for PIR.

**Table 2.** Survey-weighted associations of socioeconomic status with DMFT and self-rated good oral health among US adults, NHANES 1999–2019. Panel A shows linear regression coefficients (β) for DMFT; Panel B shows odds ratios (OR) from logistic regression models for self-rated good oral health. All models are adjusted for age, sex, race/ethnicity and additional covariates. PIR, poverty-income ratio; DMFT, decayed, missing and filled teeth.

| Panel | Predictor | Overall | Age<br>20-44 | Age<br>45-64 | Age >=65 |
| --- | --- | --- | --- | --- | --- |
| Panel A: DMFT | PIR (per 1-unit increase) | -0.60<br>(-0.64,<br>-0.57) | -0.25<br>(-0.29,<br>-0.22) | -0.79<br>(-0.87,<br>-0.72) | -1.07<br>(-1.19,<br>-0.95) |
| Panel A: DMFT | High school or GED (ref: <HS) | 0.71<br>(0.47,<br>0.95) | 0.33<br>(0.17,<br>0.50) | 0.45<br>(0.15,<br>0.76) | 1.18 (0.83,<br>1.54) |
| Panel A: DMFT | Some college | 1.16 | 0.41 | 0.99 | 1.54 (1.18, |
|  |  | (0.93,<br>1.39) | (0.26,<br>0.57) | (0.69,<br>1.29) | 1.90) |
| Panel A: DMFT | College or<br>higher | 1.74<br>(1.51,<br>1.97) | 0.43<br>(0.27,<br>0.59) | 1.90<br>(1.57,<br>2.22) | 2.59 (2.20,<br>2.98) |
| Panel B:<br>Self-rated good<br>oral health (OR) | PIR (per 1-unit<br>increase) | 1.42<br>(1.42,<br>1.42) | 1.48<br>(1.48,<br>1.48) | 1.46<br>(1.46,<br>1.46) | 1.35 (1.35,<br>1.35) |
| Panel B:<br>Self-rated good<br>oral health (OR) | High school or<br>GED (ref:<br><HS) | 1.27<br>(1.27,<br>1.27) | 1.33<br>(1.32,<br>1.33) | 1.07<br>(1.06,<br>1.07) | 1.28 (1.28,<br>1.28) |
| Panel B:<br>Self-rated good<br>oral health (OR) | Some college | 1.59<br>(1.58,<br>1.59) | 1.79<br>(1.79,<br>1.80) | 1.50<br>(1.50,<br>1.50) | 1.18 (1.18,<br>1.18) |
| Panel B:<br>Self-rated good<br>oral health (OR) | College or<br>higher | 3.45<br>(3.45,<br>3.46) | 4.71<br>(4.70,<br>4.71) | 3.09<br>(3.09,<br>3.10) | 2.01 (2.00,<br>2.01) |

Model-based predicted probabilities illustrated a clear and monotonic SES gradient across PIR quartiles within each age group (Figure 1). Among adults aged 20–44 years, the predicted probability of reporting good oral health increased from roughly 0.60 in the lowest PIR quartile to about 0.88 in the highest. Among adults aged ≥65 years, the corresponding increase was from approximately 0.67 to 0.86. Adults aged 45–64 years showed intermediate levels but a similarly steep gradient. Exploratory models including explicit SES-by-age interaction terms did not materially change these patterns beyond what was captured by age stratification.

**Figure 1.**
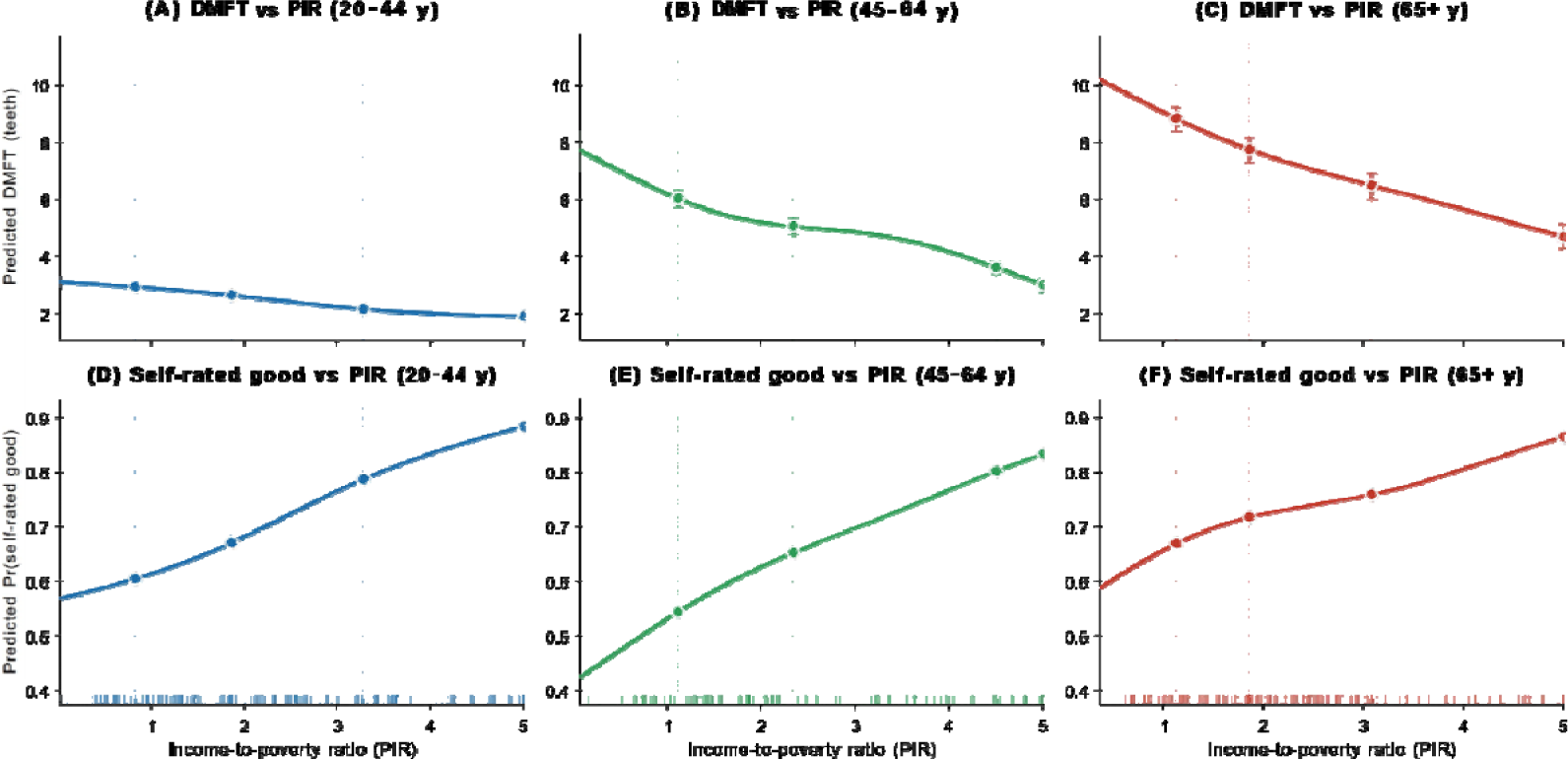
Income-to-poverty ratio gradients in oral health by age group, NHANES 1999–2019. Panels (A)–(C) show model-based predicted mean DMFT across the poverty-income ratio (PIR) distribution for adults aged 20–44, 45–64 and ≥65 years, respectively. Panels (D)–(F) show the corresponding predicted probabilities of self-rated good oral health for the same age groups. Predictions are from survey-weighted regression models including PIR modelled with restricted cubic splines (4 degrees of freedom) and adjusted for education, sex, race/ethnicity and other covariates in the main specification. Solid lines represent predicted values and shaded bands 95% confidence intervals obtained from 400 parametric draws. Vertical dashed lines indicate quartiles of the PIR distribution; rug plots along the x-axis show the distribution of PIR values in the pooled sample.

### 3.3 Unmet dental need and reasons for non-use

Unmet dental need displayed not only a pronounced income gradient but also distinct patterns in reported reasons for not obtaining care (Figure 2). Across cycles, roughly one quarter to one third of adults in the lowest PIR quartile reported unmet dental need, compared with around one in ten or fewer in the highest quartile.

**Figure 2.**
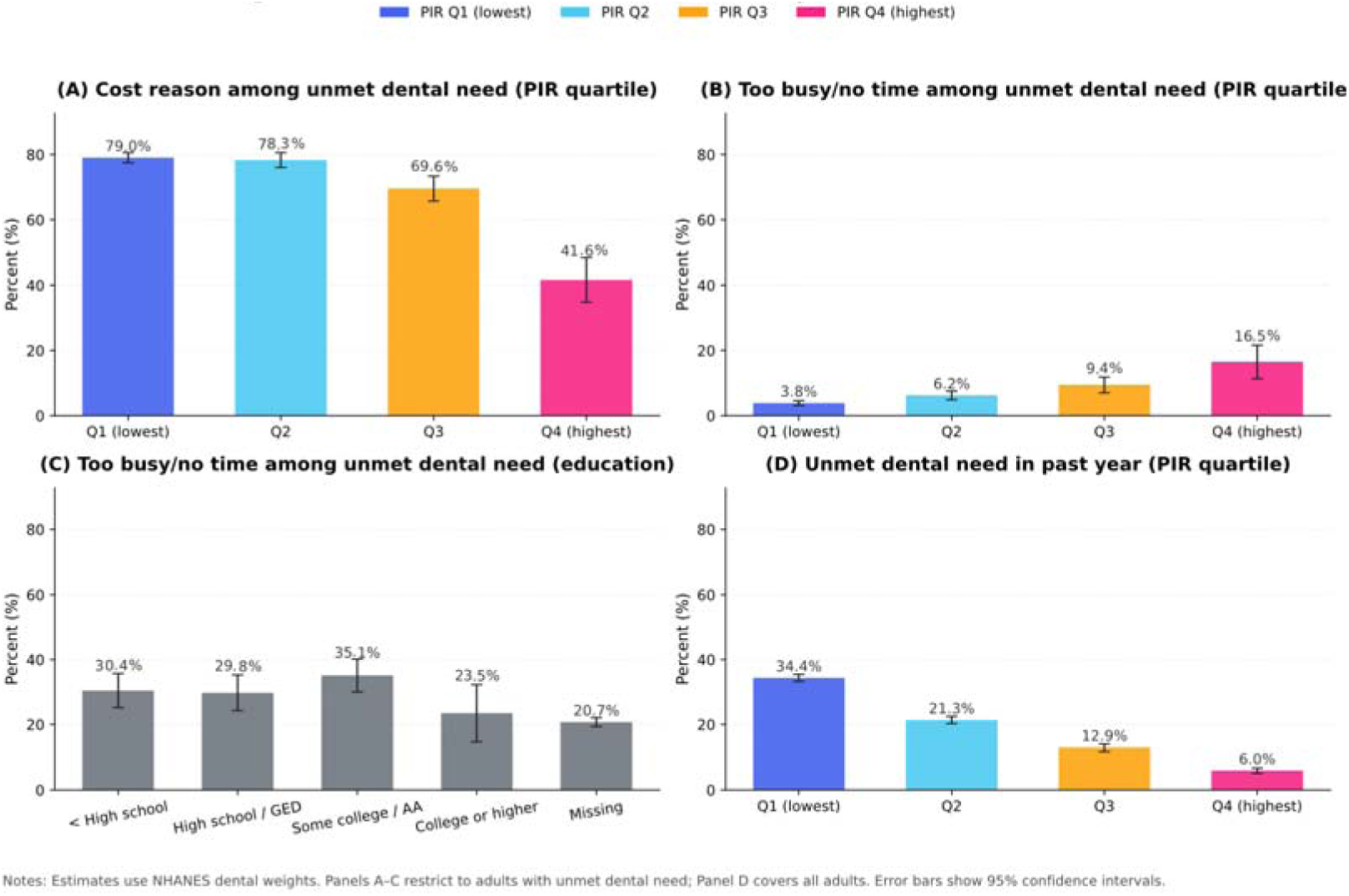
Unmet dental need and reasons for not obtaining dental care among US adults, by income and education, NHANES 2003–2019. (A) Proportion of adults with unmet dental need who cited cost as the main reason, by poverty-income ratio (PIR) quartile. (B) Proportion who cited being “too busy / no time” as the main reason, by PIR quartile. (C) The proportion of ‘not covered by insurance’ and ensure valid estimates are obtained by educational group (D) Prevalence of unmet dental need in the past year, by PIR quartile. All estimates are survey-weighted and plotted by NHANES 2-year cycles or pooled across cycles as appropriate.

Among adults who reported unmet dental need, cost was the dominant barrier in all income groups but was especially prevalent among those with low income. In the lowest PIR quartile, approximately three quarters or more (typically 75–85%) of those with unmet need cited “could not afford cost” as the main reason, with similarly high proportions in the second and third quartiles. In contrast, among adults in the highest income quartile with unmet need, cost remained the single most frequently cited reason but accounted for a smaller share, generally around one half to two thirds of cases.

Reporting being “too busy” or having “no time” as the main reason for unmet dental need was less common overall but showed the opposite socioeconomic pattern. This reason was least frequent among adults in the lowest PIR quartile, generally below 10% of those with unmet need, and more frequent among adults in the highest quartile, where it reached approximately 10–18% across cycles. A similar pattern was observed by education level: adults with college or higher education were more likely to report time constraints as the main barrier than those with less education, despite having lower overall levels of unmet need. These findings suggest that low-income adults face a high prevalence of unmet dental need that is predominantly driven by financial barriers, whereas among higher-income and higher-education adults, residual unmet need more often reflects time and organisational constraints.

### 3.4 Income-related inequality indices and decomposition

Income-related inequality indices confirmed that oral health in the United States is strongly structured by SES (Table 3). The concentration index (CI) for DMFT was approximately −0.105 overall, indicating that higher caries and tooth loss burden was concentrated among lower-income adults. By contrast, the CI for self-rated good oral health was about +0.094, indicating that favourable oral health was concentrated among higher-income adults. When stratified by age group, CIs suggested that income-related inequalities were present across the adult life course and were particularly pronounced among adults aged 45–64 years.

**Table 3.**
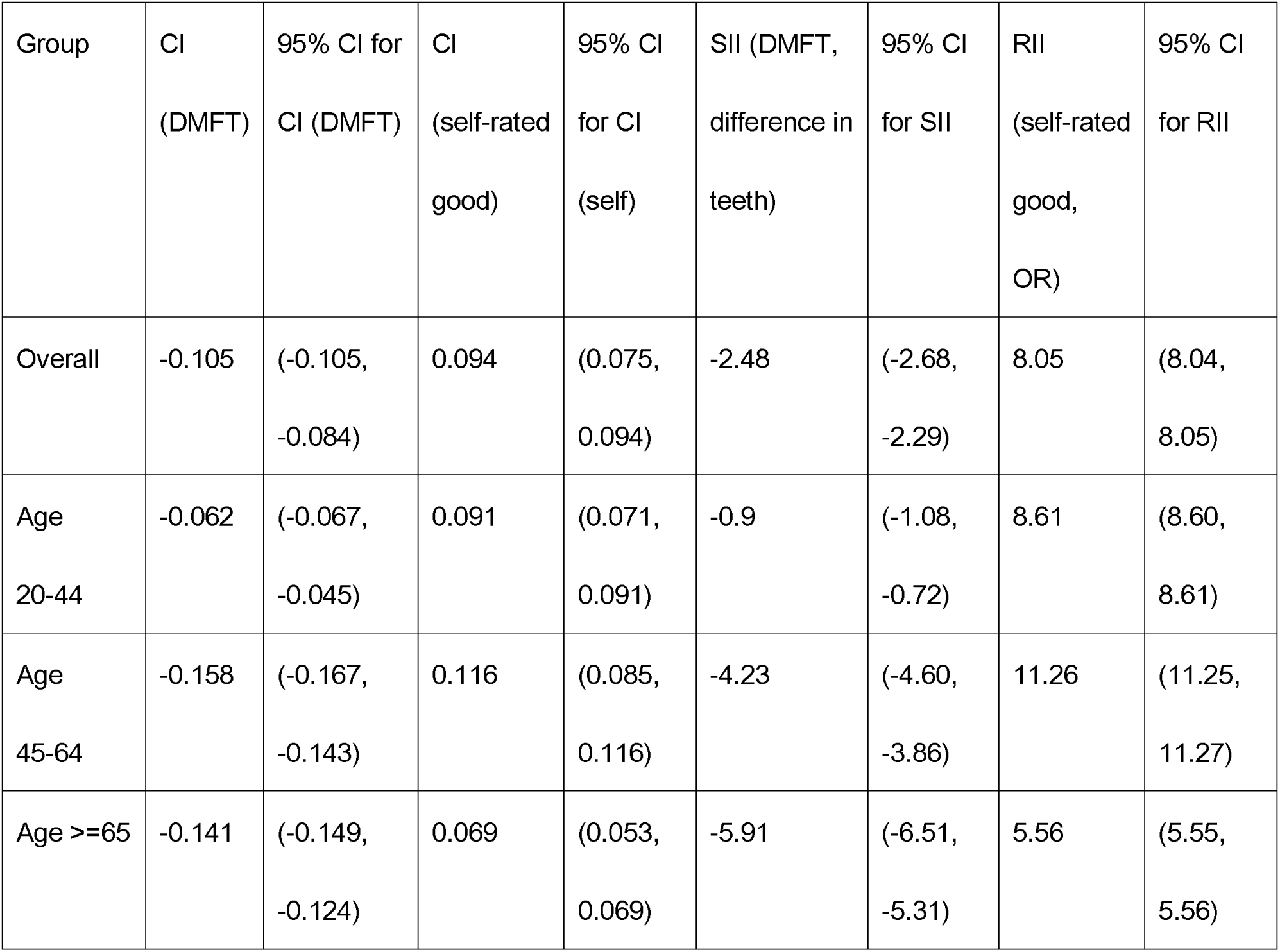
Income-related inequalities in DMFT and self-rated good oral health among US adults, overall and by age group. CI, concentration index; SII, Slope Index of Inequality; RII, Relative Index of Inequality. CI <0 for DMFT indicates concentration of disease among poorer adults; CI >0 for self-rated good oral health indicates concentration among richer adults. SII represents the absolute difference in DMFT between the lowest and highest positions in the income hierarchy; RII is the odds ratio for self-rated good oral health comparing the highest versus lowest positions.

| Group | CI<br>(DMFT) | 95% CI for<br>CI (DMFT) | CI<br>(self-rated<br>good) | 95% CI<br>for CI<br>(self) | SII (DMFT,<br>difference in<br>teeth) | 95% CI<br>for SII | RII<br>(self-rated<br>good,<br>OR) | 95% CI<br>for RII |
| --- | --- | --- | --- | --- | --- | --- | --- | --- |
| Overall | -0.105 | (-0.105,<br>-0.084) | 0.094 | (0.075,<br>0.094) | -2.48 | (-2.68,<br>-2.29) | 8.05 | (8.04,<br>8.05) |
| Age<br>20-44 | -0.062 | (-0.067,<br>-0.045) | 0.091 | (0.071,<br>0.091) | -0.9 | (-1.08,<br>-0.72) | 8.61 | (8.60,<br>8.61) |
| Age<br>45-64 | -0.158 | (-0.167,<br>-0.143) | 0.116 | (0.085,<br>0.116) | -4.23 | (-4.60,<br>-3.86) | 11.26 | (11.25,<br>11.27) |
| Age >=65 | -0.141 | (-0.149,<br>-0.124) | 0.069 | (0.053,<br>0.069) | -5.91 | (-6.51,<br>-5.31) | 5.56 | (5.55,<br>5.56) |

The Slope Index of Inequality (SII) and Relative Index of Inequality (RII) highlighted the absolute and relative magnitude of these differences. The SII for DMFT indicated that moving from the bottom to the top of the PIR rank distribution was associated with an average reduction of about 2.48 affected teeth, after adjusting for age, sex and race/ethnicity. The RII for self-rated good oral health, expressed as an odds ratio, was approximately 8.05, implying that adults at the top of the income distribution had roughly eight-fold higher odds of reporting good oral health than those at the bottom. Across age groups, SIIs and RIIs pointed to substantial inequalities, with the largest relative differences typically observed in midlife.

Mediation analysis and Oaxaca–Blinder decomposition provided further insight into pathways underpinning these gradients (Table 4). Overall, adults in the highest PIR quartile had on average 1.31 fewer affected teeth than those in the lowest quartile. The Oaxaca–Blinder decomposition attributed about 24.7% of this DMFT gap to differences in observed characteristics (“explained” component) and 75.3% to differences in coefficients and unobserved factors (“unexplained” component. Within the explained component, access-related variables—particularly having had a dental visit in the past year and not reporting unmet dental need—contributed approximately −0.88 DMFT, accounting for the largest share of the explained gap. Age (per 10-year increase) contributed a positive 0.55, partially offsetting the access effect, and race/ethnicity variables collectively contributed roughly −0.11, indicating a modest narrowing of the income gap.

**Table 4.** Oaxaca–Blinder decomposition of the difference in mean DMFT between adults in the lowest and highest PIR quartiles, NHANES 1999–2019. Panel A shows the overall Q4–Q1 difference in mean DMFT and its partition into explained and unexplained components. Panel B shows contributions of domains of explanatory variables (access to dental care, behaviour and sleep, education, age/sex and race/ethnicity) to the total gap. PIR, poverty-income ratio; DMFT, decayed, missing and filled teeth.

| Statistic(panel A) | Estimate | % of total |
| --- | --- | --- |
| Mean DMFT Q1 | 4.47 |  |
| Mean DMFT Q4 | 3.15 |  |
| Difference (Q4 - Q1) | -1.31 | 100.0 |
| Explained component | -0.28 | 21.4 |
| Unexplained component | -1.03 | 78.6 |
| Domain | Contribution to DMFT gap ($\Delta$ teeth) | % of total gap |
| Access to dental care | -0.80 | 60.7 |
| Behaviour and sleep | -0.06 | 4.3 |
| Education | 0.16 | -12.2 |
| Age and sex | 0.53 | -40.6 |
| Race/ethnicity | -0.12 | 9.2 |

In complementary mediation models, annual dental visiting, unmet dental need and short sleep duration were treated as potential mediators of the association between SES and oral health. These models were consistent with a partial indirect pathway from SES to both DMFT and self-rated oral health through dental access and sleep, with statistically significant indirect effects whose 95% confidence intervals did not cross zero. Given the cross-sectional nature of the data, these mediation estimates should be interpreted as exploratory. Taken together, the decomposition and mediation analyses highlight differential access to dental care as an important proximal pathway through which income is related to cumulative oral disease, while also indicating that a substantial share of the SES gap remains unexplained by the measured mediators and likely reflects deeper structural and life-course determinants.

### 3.5 State-level Medicaid policies and BRFSS outcomes

In the state-level difference-in-differences analysis linking the Medicaid policy panel with BRFSS outcomes, we did not detect large, precisely estimated average effects of Medicaid expansion or adult Medicaid dental benefit generosity on aggregate oral health indicators (Table 5). For the primary outcome, the state-level past-year dental visit rate, the coefficient on the Medicaid expansion indicator was 0.214 (standard error [SE] 0.0352; p=0.26) and the coefficient on the ordinal measure of adult dental benefit generosity was −0.0418 (SE 0.0378; p=0.27), with an R² of 0.219.

**Table 5.** Difference-in-differences estimates of the association between Medicaid expansion, adult Medicaid dental benefit generosity and state-level oral health indicators, BRFSS 2011–2025. Each panel reports coefficients (β), standard errors (SE) and p values from state- and year-fixed effects models with standard errors clustered at the state level. The primary outcome is the past-year dental visit rate among adults; any permanent tooth loss and complete edentulism among adults aged ≥65 years are examined as exploratory outcomes

| Outcome | Variable | Beta | SE | p |
| --- | --- | --- | --- | --- |
| dental_visit_rate | MedicaidExpansion_st | 0.214 | 0.878 | 0.807 |
| dental_visit_rate | DentalGenerosityLevel_st | -0.373 | 0.950 | 0.695 |
| any_tooth_loss_rate | MedicaidExpansion_st | -0.001 | 0.002 | 0.591 |
| any_tooth_loss_rate | DentalGenerosityLevel_st | -0.002 | 0.002 | 0.273 |
| edentulism_65_rate | MedicaidExpansion_st | -2.903 | 1.755 | 0.098 |
| edentulism_65_rate | DentalGenerosityLevel_st | -0.761 | 5.452 | 0.889 |
| dental_visit_rate | R<sup>2</sup> | 0.300 |  |  |
| any_tooth_loss_rate | R<sup>2</sup> | 0.209 |  |  |
| edentulism_65_rate | R<sup>2</sup> | 0.434 |  |  |

For any permanent tooth loss, Medicaid expansion was associated with a change of −0.012 in prevalence (SE 0.034; p=0.73), and adult dental benefit generosity with a small, non-significant positive coefficient of about +0.008. For complete edentulism among adults aged ≥65 years, the coefficients were +0.241 (SE 0.334; p=0.47) for Medicaid expansion and approximately −0.125 for dental benefit generosity, neither statistically significant. Graphical inspection of state-level time trends in dental visit rates, any tooth loss and edentulism showed broadly parallel trajectories for expansion versus non-expansion states and for states with more versus less generous adult dental benefits, with no consistent evidence of divergence following policy changes (Figure 3). These results suggest that, over the study period examined, changes in Medicaid eligibility and adult dental benefit generosity, as measured here, were not associated with large changes in state-wide averages of dental visits or tooth loss, although effects in specific low-income subgroups cannot be ruled out.

**Figure 3.**
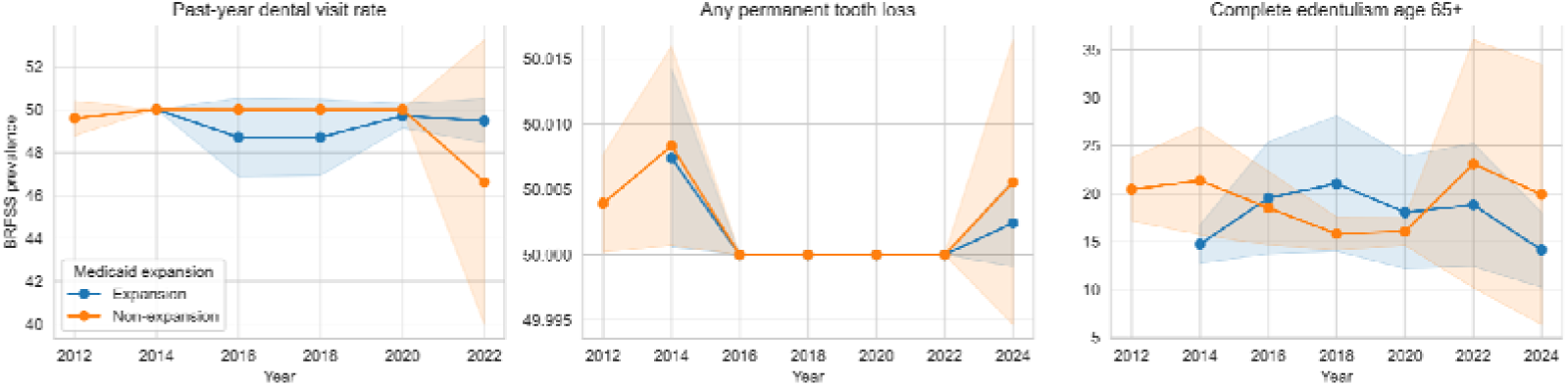
Trends in state-level oral health indicators by Medicaid expansion status, BRFSS 2011–2025. (A) Past-year dental visit rate among adults. (B) Prevalence of any permanent tooth loss among adults. (C) Prevalence of complete edentulism among adults aged ≥65 years. Lines show mean age-adjusted prevalence in states that adopted the Affordable Care Act Medicaid expansion versus those that did not; shaded areas or error bars (if shown) represent 95% confidence intervals.

### 3.6 Macro-level WHO patterns

Descriptive analyses of WHO country-level data confirmed substantial geographical variation in the burden of oral disease and in dental service capacity. Countries and regions with higher national income and health expenditure tended to have lower age-standardised prevalence of severe periodontal disease and edentulism, but these associations were neither linear nor uniform and were sensitive to population age structure and data quality. In some settings, higher per capita dental expenditure coincided with higher reported edentulism and greater use of prosthetic treatments, reflecting cumulative historical disease and differences in treatment preferences and reporting practices. These macro-level patterns highlight the complexity of relationships between economic development, health systems and oral health at the country level, and underscore the value of individual-level analyses in clarifying the pathways through which socioeconomic status shapes oral health inequalities.

## 4 Discussion

### 4.1 Principal findings

In this nationally representative analysis of US adults, we found strong and consistent socioeconomic gradients in both objective and subjective oral health. Higher income and higher education were associated with fewer decayed, missing and filled teeth (DMFT) and higher probabilities of self-rated good oral health across the adult life course. Inequality indices showed that DMFT burden was concentrated among lower-income adults, whereas good self-rated oral health was concentrated among those with higher income, with large absolute and relative differences between the bottom and top of the income distribution.

We further showed that these gradients are closely linked to access to dental care. Low-income adults had a high prevalence of unmet dental need, and most unmet need in this group was attributed to inability to afford care. Decomposition and mediation analyses indicated that differences in annual dental visiting and unmet dental need explained a substantial share of the income gap in cumulative caries experience, although a large proportion of the gap remained unexplained. Sleep-related behaviours also contributed, but to a lesser extent.

By contrast, state-level difference-in-differences models did not reveal large, precisely estimated average effects of Medicaid expansion or adult Medicaid dental benefit generosity on aggregate state-level dental visit rates, tooth loss or edentulism among older adults over the study period examined. Macro-level analyses of WHO data confirmed large geographical variation in oral disease burden and dental service capacity and highlighted complex, non-linear associations between national income, dental expenditure and oral health outcomes. Taken together, these findings suggest that oral health inequalities in the United States are substantial, that individual-level socioeconomic position and access to dental care remain central determinants, and that recent changes in Medicaid eligibility and adult dental benefits, while important, may not be sufficient on their own to shift distal, population-level oral health indicators.

### 4.2 Interpretation in light of mechanisms and previous evidence

Our results are consistent with social determinants frameworks in which material, behavioural, psychosocial and institutional pathways jointly link socioeconomic status (SES) to oral health. Material constraints are evident in the high prevalence of cost-related unmet dental need among low-income adults[23]. Despite similar levels of perceived need, adults in the lowest income quartile were several times more likely than those in the highest quartile to report needing dental care but not obtaining it, and cost was overwhelmingly cited as the main barrier in this group. This pattern is aligned with previous NHANES-based studies and national surveys reporting that financial barriers remain the dominant reason for forgone dental care among socioeconomically disadvantaged US adults.

Behavioural and sleep-related pathways also appear to play a role. Higher income and education are typically associated with healthier behaviours, including more regular toothbrushing with fluoride toothpaste, lower intake of sugary foods and beverages, lower smoking prevalence and more adequate sleep. In our analyses, short sleep duration and other behavioural indicators showed independent associations with oral health and mediated part of the SES–oral health relationship, although their contribution was smaller than that of dental access variables. This finding is compatible with broader evidence linking insufficient sleep to systemic inflammation, immune dysregulation and increased risk of both chronic disease and poorer self-rated health, including oral health.

Psychosocial stress and structural conditions are more difficult to observe directly but are likely to underlie some of the unexplained component in the Oaxaca–Blinder decomposition[24, 25]. The substantial “unexplained” share of the income–DMFT gap suggests that differences in coefficients and unmeasured factors—such as chronic stress, precarious employment, neighbourhood conditions, early-life exposures, and experiences of discrimination in health care and labour markets—contribute meaningfully to oral health inequalities[9, 26]. Racial and ethnic differences in oral health, which persist even after adjusting for income and education in many studies, are consistent with the impact of long-standing structural and institutional disadvantages that shape both SES and health among minoritised groups.

At the institutional level, our findings highlight the central role of health systems in structuring access to care. Oral health services for adults in the United States are predominantly financed through private dental insurance and out-of-pocket payments, with public coverage largely limited to Medicaid and highly variable across states. Our NHANES analyses show that regular dental visiting is strongly patterned by SES, and our decomposition results indicate that differences in dental access account for a large portion of the SES gap in DMFT. Yet the state-level analyses suggest that recent changes in Medicaid expansion and adult dental benefit generosity have not translated into large shifts in state-wide averages of dental visits, tooth loss or edentulism over the follow-up period.

Several explanations are plausible. First, tooth loss and edentulism are highly cumulative outcomes that reflect lifetime disease and treatment histories; reforms concentrated in the 2010s may require longer time horizons to affect these distal indicators. Second, BRFSS outcomes reflect state-wide prevalence in the entire adult population, including many individuals who are not Medicaid-eligible; any improvements confined to low-income enrollees may therefore be diluted when aggregated to the state level. Third, our policy indicators, although based on established sources, cannot fully capture differences in benefit scope, reimbursement rates, provider participation and implementation intensity across states. Previous individual-level studies have reported that Medicaid expansion and enhanced adult dental benefits can increase dental visiting and reduce cost-related unmet need among low-income adults. Our state-level results do not contradict these findings but suggest that, in their current form and timeframe, such policies have limited capacity to alter broad, population-level averages in cumulative oral disease.

International comparisons provide an additional lens for interpreting these findings. National oral health surveys in China, for example, have documented substantial urban–rural, regional and socioeconomic disparities in dental caries, periodontal disease and tooth loss, despite improvements in average indicators and the expansion of basic medical insurance[4, 27–29]. Similar to the United States, adult dental services in China are only partially covered, with preventive and restorative care often requiring substantial out-of-pocket spending. Cross-country WHO data suggest that high-income countries with more comprehensive dental coverage and lower out-of-pocket costs tend to have better average oral health, but pronounced social gradients often persist within countries. Together with our results, this evidence indicates that expanding insurance coverage and service availability is necessary but not sufficient to eliminate oral health inequalities; underlying income, education and broader social and structural determinants must also be addressed.

### 4.3 Policy implications

The contrast between strong individual-level SES gradients and modest, imprecise state-level policy effects has important implications for oral health policy in the United States and for countries such as China that are seeking to integrate oral health into universal health coverage agendas.

First, our findings underscore the importance of reducing financial barriers to dental care for low-income adults. The very high prevalence of cost-related unmet dental need in the lowest income quartile suggests that expanding publicly financed dental coverage remains a central priority. In the US context, this includes extending and stabilising adult dental benefits in Medicaid, moving towards more comprehensive coverage of preventive and restorative services, and improving reimbursement and administrative conditions to support provider participation. Similar arguments apply to other publicly financed schemes, including Medicare and state or local safety-net programmes [23].

Second, the persistent SES gradients and the large residual “unexplained” component after accounting for access and key behaviours indicate that broader social and economic policies will be needed to narrow oral health inequalities. Policies that reduce poverty, improve wages and working conditions, and expand educational opportunities are likely to yield downstream benefits for oral as well as general health. Interventions that address time-related and organisational barriers—such as flexible clinic hours, workplace policies that allow paid leave for dental visits, and integration of basic oral health services into primary care and community health settings—may be particularly relevant for higher-income and higher-education groups whose residual unmet need is more often driven by time constraints than by inability to pay.

Third, our results support the view that oral health policy cannot be separated from broader health system design. Decisions about whether and how dental care is included in basic benefit packages, the balance between public and private financing, the level of cost-sharing, and the organisation of primary care all influence the distribution of oral health. For China and other countries expanding universal health coverage, incorporating a meaningful set of adult dental services into public benefits, strengthening community-based preventive care, and aligning fiscal and workforce policies with oral health goals may help avoid reproducing the wide inequalities observed in settings where adult dental care remains largely market-based.

Finally, the interplay between SES, race/ethnicity and oral health points to the need for policies that explicitly address structural and institutional determinants. This includes tackling residential segregation and provider maldistribution, ensuring equitable access to high-quality dental care in underserved communities, and monitoring the distributional impact of policies across socioeconomic and racial/ethnic groups. Without attention to these upstream determinants, there is a risk that new benefits and technologies will be taken up preferentially by more advantaged groups, potentially widening rather than narrowing oral health gaps.

### 4.4 Strengths and limitations

This study has several strengths. We used large, nationally representative datasets with standardised clinical and self-reported measures of oral health, allowing us to characterise socioeconomic gradients across the adult life course. We applied multiple complementary measures of inequality (CI, SII and RII), which provide both absolute and relative perspectives on income-related differences. By combining mediation analysis and Oaxaca–Blinder decomposition, we moved beyond description to examine potential mechanisms, quantifying the contribution of access to dental care and selected behaviours to socioeconomic gaps in cumulative oral disease. The integration of individual-level NHANES analyses with a state-level Medicaid policy panel and BRFSS outcomes is another strength, enabling us to relate individual-level inequalities to broader policy changes. Finally, the use of WHO country-level data provides international context for interpreting US findings.

Several limitations should also be acknowledged. First, the NHANES analyses are cross-sectional and cannot establish causal effects of SES, access or behaviours on oral health. Reverse causation and residual confounding are possible, although sensitivity analyses adjusting for additional covariates yielded similar patterns. Second, mediation and decomposition analyses rely on assumptions about model specification and the absence of unmeasured confounding of the mediator–outcome relationships; violations of these assumptions could bias estimates of indirect effects and domain contributions. Third, BRFSS outcomes are based on self-reported prevalence and reflect aggregate state-level patterns; they may underestimate policy effects confined to specific subgroups, such as low-income Medicaid beneficiaries. Our Medicaid policy indicators, although drawn from authoritative sources, cannot fully capture within-state variation in benefit scope, reimbursement levels, provider participation and implementation over time. Fourth, WHO country-level data are subject to heterogeneity in data quality and reporting practices, limiting the precision of cross-country comparisons.

Finally, our study focuses on income and education as core SES indicators and on a limited set of mediators. Other dimensions of social position, such as wealth, occupation, early-life circumstances and neighbourhood conditions, and other potential mediators, including dietary patterns, oral hygiene practices and psychosocial factors, were not fully explored and warrant further investigation.

### 4.5 Conclusions

In summary, we find that oral health inequalities among US adults are large, persistent and strongly patterned by socioeconomic position. Higher income and education are associated with fewer decayed, missing and filled teeth and better self-rated oral health, and income-related inequalities are substantial in both absolute and relative terms. Differences in access to dental care—particularly regular dental visiting and the burden of unmet dental need—account for an important part of these gradients, but a sizeable share remains unexplained, pointing to deeper structural determinants. Recent Medicaid expansions and enhancements in adult dental benefits, while important for improving financial protection for low-income adults, have not yet translated into large changes in aggregate state-level indicators of dental visiting or tooth loss.

Efforts to reduce oral health inequalities will need to combine expansion and strengthening of publicly financed dental coverage with broader policies that address income and education inequalities, structural and institutional barriers, and the social patterning of health behaviours. Lessons from the United States are relevant for China and other countries seeking to incorporate oral health into universal health coverage: without confronting upstream social and economic determinants, gains in average oral health are likely to coexist with, and may even obscure, substantial and persistent inequalities.

## Data Availability

All data produced in the present study are available upon reasonable request to the authors

https://www.cdc.gov/nchs/nhanes/index.html

https://www.kaggle.com/datasets/luciadam/oral-health-quire

https://www.chcs.org/media/Medicaid-Adult-Dental-Benefits-Overview-Appendix_091519.pdf

https://www.carequest.org/Medicaid-Adult-Dental-Coverage-Checker

https://www.macpac.gov/subtopic/medicaid-expansion/

