## Supplementary Material for "Income, access to care and adult oral health inequalities in the United States: a multilevel analysis of national surveys and Medicaid policies"

Zhou Chengke[1]

[1] The State Key Laboratory of Oral & Maxillofacial Reconstruction and Regeneration, Key Laboratory of Oral Biomedicine Ministry of Education, Hubei Key Laboratory of Stomatology, School & Hospital of Stomatology, Frontier Science Center for Immunology and Metabolism, Taikang Center for Life and Medical Sciences, Wuhan University, Wuhan, 430079, China.

### Appendix. Detailed statistical methods

#### A1. Survey design and weighting

All NHANES analyses used the complex, multistage probability design of the continuous survey from 1999 onwards. We combined adults aged ⩾20 years from successive 2-year cycles, restricting to participants with completed oral examinations and non-missing values for key variables. Following NCHS analytic guidelines, we used the examination sample weights appropriate for multi-cycle analyses, along with strata and primary sampling units (PSUs), and implemented Taylor series linearization for variance estimation. When pooling multiple cycles, weights were divided by the number of combined cycles so that the sum of weights represented the average annual non-institutionalised US population over the study period, as recommended in NHANES analytic guidance.

For state-level analyses, we used BRFSS annual data from participating US states and the final raked weights provided by CDC. BRFSS employs random-digit-dial sampling, combining landline and cellular telephones, with iterative proportional fitting (raking) to align the sample with state-level population benchmarks on age, sex, race/ethnicity, education and other factors. All prevalence estimates and regressions used the final BRFSS weights and design variables, with variance estimated using Taylor series methods.

#### A2. Socioeconomic ranking and ridit scores

Socioeconomic status was measured using the family poverty–income ratio (PIR), defined by NHANES as the ratio of family income to the federal poverty threshold for that year. PIR was analysed both as a continuous variable and as quartiles (Q1–Q4). For inequality indices, we used the individual’s relative rank in the income distribution, denoted $r_i$, obtained by ordering adults by PIR and assigning each individual the mid-point of their cumulative population share (ridit score). In a sample of size $n$, the ridit for individual $i$with rank $R_i$(1 = poorest, $n$= richest) is:

$$r_i = \frac{2R_i - 1}{2n}$$

so that $r_i$ranges from approximately 0 (lowest PIR) to 1 (highest PIR). When survey weights were used, ranks were computed based on weighted cumulative distributions. Similar ridit scores were constructed for ordered education categories when computing education-related inequalities.

#### A3. Concentration index for income-related inequality

We quantified income-related inequality in oral health using the concentration index (CI), which measures the degree to which a health variable is concentrated among richer versus poorer individuals.

Let $y_i$denote the health variable (e.g. DMFT or an indicator of good self-rated oral health) for individual $i$, with mean $\mu$, and let $r_i$be the individual's fractional income rank. The standard (relative) concentration index is defined as:

$$C = \frac{2}{\mu} \operatorname{cov}(y_i, r_i),$$

where the covariance is taken over individuals in the population (or sample, using survey weights). $C$ranges from −1 to +1. A negative CI for an adverse outcome (e.g. DMFT) indicates that the burden is disproportionately concentrated among the poor; a positive CI for a favourable outcome (e.g. self-rated good oral health) indicates concentration among the better-off.

In practice, we estimated $C$using a regression-based convenient formula with survey weights, which is algebraically equivalent to the covariance definition, and used linearization methods to derive standard errors and 95% confidence intervals, following standard World Bank and WHO guidance.

#### A4. Slope index of inequality (SII) and relative index of inequality (RII)

To complement CI, we computed absolute and relative inequality measures using the slope index of inequality (SII) and relative index of inequality (RII).

For each ordered socioeconomic variable (income or education), individuals were assigned a ridit score $r_i$ as described above. For a continuous oral health outcome such as DMFT, we fitted a survey-weighted linear regression:

$$y_i = \alpha + \beta r_i + \epsilon_i,$$

where $y_i$ is the outcome and $r_i$ is the socioeconomic rank. The SII is defined as the difference in predicted outcome between the top and bottom of the socioeconomic distribution:

$$\text{SII} = \hat{y}(r = 1) - \hat{y}(r = 0) = \hat{\beta},$$

so that a negative SII for DMFT indicates fewer affected teeth among those at the top of the income distribution compared with those at the bottom.

For binary outcomes, such as "self-rated good oral health", we fitted generalized linear models with a log or logit link on the same rank variable and covariates. For models with a log link, the RII is defined as the ratio of predicted probabilities:

$$\text{RII} = \frac{\hat{p}(r = 1)}{\hat{p}(r = 0)},$$

where $\hat{p}(r)$ is the predicted probability of the outcome at rank $r$. An RII greater than 1 indicates that the favourable outcome is more common among the highest socioeconomic group. Where logistic models were used, we report RII on the odds scale, calculated as the ratio of predicted odds at $r = 1$ versus $r = 0$. Confidence intervals for SII and RII were obtained from the model-based standard errors under the survey design.

#### A5. Oaxaca–Blinder decomposition of income-related differences in DMFT

To separate the extent to which income differences in DMFT are attributable to differences in observed characteristics versus differences in the effects of those characteristics, we used a twofold Oaxaca–Blinder decomposition.

We considered two groups: adults in the lowest PIR quartile (Q1) and those in the highest quartile (Q4). Let $\bar{y}_1$and $\bar{y}_4$denote the mean DMFT in Q1 and Q4, respectively, and let $\bar{X}_1$and $\bar{X}_4$be the vectors of mean covariate values in each group, including access variables (recent dental visit, unmet dental need), sleep duration, education indicators, age (in 10-year units), sex and race/ethnicity. We estimated separate survey-weighted linear regressions for each group:

$$y_{gi} = X_{gi}^{\top}\beta_g + \epsilon_{gi}, g \in \{1,4\},$$

and a pooled regression in the combined sample to obtain a reference coefficient vector $\beta^*$. The difference in mean DMFT between Q4 and Q1 can be written as:

$$\Delta = \bar{y}_4 - \bar{y}_1 = (\bar{X}_4 - \bar{X}_1)^{\top}\beta^* \ + \ \bar{X}_4^{\top}(\beta_4 - \beta^*) \ + \ \bar{X}_1^{\top}(\beta^* - \beta_1).$$

The first term, $(\bar{X}_4 - \bar{X}_1)^{\top}\beta^*$, is the “explained” component, representing the part of the gap that would remain if the two groups shared the same coefficients $\beta^*$but differed in their covariate distributions. The remaining terms are combined as the “unexplained” component, capturing differences in regression coefficients and unmeasured factors.

For interpretability, we grouped covariates into domains (access, behaviours/sleep, education, age/sex, race/ethnicity) and summed their contributions within the explained component. A positive domain contribution indicates that differences in that domain widen the Q4–Q1 DMFT gap, whereas a negative contribution indicates that they narrow the gap. Standard errors for decomposition components were obtained using the delta method implemented in standard Oaxaca–Blinder routines adapted for survey data.

#### A6. Mediation analysis

To explore potential pathways linking socioeconomic status to oral health, we conducted mediation analyses treating dental access and sleep as potential mediators, using product-of-coefficients approaches with bootstrap confidence intervals.

In the simplest form, let $X$ denote the socioeconomic exposure (continuous PIR or education), $M$ a mediator (e.g. any dental visit in the past year, unmet dental need, short sleep duration) and $Y$ an outcome (DMFT or self-rated good oral health). For each mediator, we fitted:

1. A mediator model:

$$M_i = \alpha_M + aX_i + \gamma^\top Z_i + \varepsilon_{Mi},$$

   where $Z_i$ is a vector of covariates (age, sex, race/ethnicity and other controls), using linear or logistic regression as appropriate.

2. An outcome model including both exposure and mediator:

$$Y_i = \alpha_Y + c'X_i + bM_i + \delta^\top Z_i + \varepsilon_{Yi},$$

   with linear models for DMFT and logistic models for self-rated good oral health.

All models were estimated with survey weights and design variables. The indirect effect via mediator $M$ was estimated as the product $\hat{a}\hat{b}$; the total effect was $\hat{c} = \widehat{c'} + \hat{a}\hat{b}$ in the linear case, and the direct effect was $\widehat{c'}$. For each mediator and outcome, we drew 300 nonparametric bootstrap samples (resampling individuals with replacement within the survey data, reapplying weights) and re-estimated $\hat{a}$ and $\hat{b}$ to obtain empirical confidence intervals for the indirect effect. When multiple mediators were included simultaneously, we estimated separate indirect effects for each mediator by extending the outcome model to include all mediators.

Because NHANES is cross-sectional, these mediation analyses rely on strong assumptions (no unmeasured confounding of exposure–mediator and mediator–outcome relations, and no exposure-induced confounders of the mediator–outcome relation) and should be interpreted as exploratory and hypothesis-generating rather than strictly causal.

##### A7. Difference-in-differences models for Medicaid expansion and dental benefits

State-level policy analyses used a difference-in-differences (DID) framework linking BRFSS outcomes to a constructed panel of state-year Medicaid expansion status and adult dental benefit generosity. For each state $s$and year $t$, we defined:

- $Y_{st}$: state-level outcome (e.g. age-adjusted dental visit rate, any tooth loss rate, edentulism among adults aged ≥65 years), estimated from BRFSS using survey weights;
- $\text{Expansion}_{st}$: indicator equal to 1 in years when Affordable Care Act Medicaid expansion was in effect in state $s$, 0 otherwise;
- $\text{DentalGenerosity}_{st}$: ordinal measure of adult Medicaid dental benefit generosity (e.g. emergency only, limited, extensive), harmonised across data sources.

We estimated linear two-way fixed effect models of the form:

$$Y_{st} = \alpha + \beta_1 \,\text{Expansion}_{st} + \beta_2 \,\text{DentalGenerosity}_{st} + \lambda_s + \delta_t + \varepsilon_{st},$$

where $\lambda_s$are state fixed effects, capturing time-invariant differences between states, and $\delta_t$are year fixed effects, capturing national shocks common to all states. In some specifications we additionally adjusted for time-varying state covariates (e.g. demographic composition) where available. Standard errors were clustered at the state level to allow for arbitrary serial correlation within states over time.

The causal interpretation of $\beta_1$and $\beta_2$as average policy effects relies on a parallel trends assumption: in the absence of policy changes, outcome trends would have been similar across states with different Medicaid expansion timing or dental benefit generosity. As a robustness check, we estimated event-study models replacing the policy indicator with a set of leads and lags relative to the year of Medicaid expansion, allowing us to inspect pre-treatment trends and the dynamic pattern of post-expansion effects. Event-study coefficients were plotted with 95% confidence intervals to assess whether pre-policy coefficients were close to zero and whether any systematic pattern emerged after expansion.

#### A8. Modelling of non-linear relationships

To allow for potential non-linear associations between continuous SES measures and outcomes, we fitted models in which PIR entered as a restricted cubic spline. In these models, PIR was transformed using a natural cubic spline basis with pre-specified knots at selected quantiles of the PIR distribution (for example, at the 5th, 35th, 65th and 95th percentiles). The outcome was then regressed on the spline basis functions and covariates using survey-weighted linear or logistic regression, and Wald tests of the joint significance of the non-linear spline terms were used to assess departures from linearity. Predicted probabilities of good self-rated oral health across PIR levels and age strata were generated from these spline models for plotting.

---

**Supplementary Tables**

**Table S1. Additional characteristics of NHANES participants by PIR quartile and education level**

| Panel | Group | Characteristic | Value | Display | Format |
|---|---|---|---|---|---|
| Panel A: By PIR quartile | Overall | Unweighted N | 60207 | 60207 | int |
| Panel A: By PIR quartile | PIR Q1 (lowest) | Unweighted N | 18829 | 18829 | int |
| Panel A: By PIR quartile | PIR Q2 | Unweighted N | 14419 | 14419 | int |
| Panel A: By PIR quartile | PIR Q3 | Unweighted N | 20887 | 20887 | int |
| Panel A: By PIR quartile | PIR Q4 (highest) | Unweighted N | 0 | 0 | int |
| Panel A: By PIR quartile | Overall | Weighted % female | 51.92862 | 51.90% | percent1 |
| Panel A: By PIR quartile | PIR Q1 (lowest) | Weighted % female | 56.70003 | 56.70% | percent1 |
| Panel A: By PIR quartile | PIR Q2 | Weighted % female | 52.28587 | 52.30% | percent1 |
| Panel A: By PIR quartile | PIR Q3 | Weighted % female | 49.19699 | 49.20% | percent1 |
| Panel A: By PIR quartile | PIR Q4 (highest) | Weighted % female | | | percent1 |
| Panel A: By PIR quartile | Overall | Mean age (years) | 47.06626 | 47.1 | mean1 |
| Panel A: By PIR quartile | PIR Q1 (lowest) | Mean age (years) | 44.2868 | 44.3 | mean1 |
| Panel A: By PIR quartile | PIR Q2 | Mean age (years) | 47.59157 | 47.6 | mean1 |
| Panel A: By PIR quartile | PIR Q3 | Mean age (years) | 47.89206 | 47.9 | mean1 |

| | | | | | |
|---|---|---|---|---|---|
| Panel A: By PIR quartile | PIR Q4 (highest) | Mean age (years) | | | mean1 |
| Panel A: By PIR quartile | Overall | BMI (kg/m^2) | 28.85003 | 28.9 | mean1 |
| Panel A: By PIR quartile | PIR Q1 (lowest) | BMI (kg/m^2) | 29.18592 | 29.2 | mean1 |
| Panel A: By PIR quartile | PIR Q2 | BMI (kg/m^2) | 29.22281 | 29.2 | mean1 |
| Panel A: By PIR quartile | PIR Q3 | BMI (kg/m^2) | 28.53634 | 28.5 | mean1 |
| Panel A: By PIR quartile | PIR Q4 (highest) | BMI (kg/m^2) | | | mean1 |
| Panel A: By PIR quartile | Overall | Current smoker (%) | 45.2652 | 45.30% | percent1 |
| Panel A: By PIR quartile | PIR Q1 (lowest) | Current smoker (%) | 51.09193 | 51.10% | percent1 |
| Panel A: By PIR quartile | PIR Q2 | Current smoker (%) | 48.6629 | 48.70% | percent1 |
| Panel A: By PIR quartile | PIR Q3 | Current smoker (%) | 41.03257 | 41.00% | percent1 |
| Panel A: By PIR quartile | PIR Q4 (highest) | Current smoker (%) | | | percent1 |
| Panel A: By PIR quartile | Overall | Sugar-sweetened drink frequency (times/day) | 2.933075 | 2.9 | mean1 |
| Panel A: By PIR quartile | PIR Q1 (lowest) | Sugar-sweetened drink frequency (times/day) | 3.1409 | 3.1 | mean1 |
| Panel A: By PIR quartile | PIR Q2 | Sugar-sweetened drink frequency (times/day) | 3.004416 | 3 | mean1 |
| Panel A: By PIR quartile | PIR Q3 | Sugar-sweetened drink frequency (times/day) | 2.791032 | 2.8 | mean1 |
| Panel A: By PIR quartile | PIR Q4 (highest) | Sugar-sweetened drink frequency (times/day) | | | mean1 |
| Panel A: By PIR quartile | Overall | Total sugar intake (g/day) | 113.7138 | 113.7 | mean1 |
| Panel A: By PIR quartile | PIR Q1 (lowest) | Total sugar intake (g/day) | 121.5297 | 121.5 | mean1 |
| Panel A: By PIR quartile | PIR Q2 | Total sugar intake (g/day) | 115.2083 | 115.2 | mean1 |
| Panel A: By PIR quartile | PIR Q3 | Total sugar intake (g/day) | 109.7414 | 109.7 | mean1 |
| Panel A: By PIR quartile | PIR Q4 (highest) | Total sugar intake (g/day) | | | mean1 |
| Panel A: By PIR quartile | Overall | Self-rated good oral health (%) | 72.24626 | 72.20% | percent1 |

| | | | | | |
|---|---|---|---|---|---|
| Panel A: By PIR quartile | PIR Q1 (lowest) | Self-rated good oral health (%) | 55.43264 | 55.40% | percent1 |
| Panel A: By PIR quartile | PIR Q2 | Self-rated good oral health (%) | 67.54395 | 67.50% | percent1 |
| Panel A: By PIR quartile | PIR Q3 | Self-rated good oral health (%) | 83.50934 | 83.50% | percent1 |
| Panel A: By PIR quartile | PIR Q4 (highest) | Self-rated good oral health (%) | | | percent1 |
| Panel A: By PIR quartile | Overall | Past-year dental visit (%) | 59.45722 | 59.50% | percent1 |
| Panel A: By PIR quartile | PIR Q1 (lowest) | Past-year dental visit (%) | 41.19872 | 41.20% | percent1 |
| Panel A: By PIR quartile | PIR Q2 | Past-year dental visit (%) | 52.54348 | 52.50% | percent1 |
| Panel A: By PIR quartile | PIR Q3 | Past-year dental visit (%) | 72.83942 | 72.80% | percent1 |
| Panel A: By PIR quartile | PIR Q4 (highest) | Past-year dental visit (%) | | | percent1 |
| Panel A: By PIR quartile | Overall | Unmet dental need (%) | 18.7667 | 18.80% | percent1 |
| Panel A: By PIR quartile | PIR Q1 (lowest) | Unmet dental need (%) | 35.00615 | 35.00% | percent1 |
| Panel A: By PIR quartile | PIR Q2 | Unmet dental need (%) | 21.94679 | 21.90% | percent1 |
| Panel A: By PIR quartile | PIR Q3 | Unmet dental need (%) | 8.689273 | 8.70% | percent1 |
| Panel A: By PIR quartile | PIR Q4 (highest) | Unmet dental need (%) | | | percent1 |
| Panel B: By education level | Overall | Unweighted N | 60207 | 60207 | int |
| Panel B: By education level | < High school | Unweighted N | 1039 | 1039 | int |
| Panel B: By education level | High school / GED | Unweighted N | 1749 | 1749 | int |
| Panel B: By education level | Some college / AA | Unweighted N | 2370 | 2370 | int |
| Panel B: By education level | College or higher | Unweighted N | 2625 | 2625 | int |
| Panel B: By education level | Missing | Unweighted N | 52424 | 52424 | int |
| Panel B: By education level | Overall | Weighted % female | 51.92862 | 51.90% | percent1 |

| | | | | | |
|---|---|---|---|---|---|
| Panel B: By education level | < High school | Weighted % female | 47.99497 | 48.00% | percent1 |
| Panel B: By education level | High school / GED | Weighted % female | 48.71659 | 48.70% | percent1 |
| Panel B: By education level | Some college / AA | Weighted % female | 53.32921 | 53.30% | percent1 |
| Panel B: By education level | College or higher | Weighted % female | 53.22191 | 53.20% | percent1 |
| Panel B: By education level | Missing | Weighted % female | 51.97355 | 52.00% | percent1 |
| Panel B: By education level | Overall | Mean age (years) | 47.06626 | 47.1 | mean1 |
| Panel B: By education level | < High school | Mean age (years) | 52.61259 | 52.6 | mean1 |
| Panel B: By education level | High school / GED | Mean age (years) | 49.27501 | 49.3 | mean1 |
| Panel B: By education level | Some college / AA | Mean age (years) | 48.0638 | 48.1 | mean1 |
| Panel B: By education level | College or higher | Mean age (years) | 47.91497 | 47.9 | mean1 |
| Panel B: By education level | Missing | Mean age (years) | 46.87094 | 46.9 | mean1 |
| Panel B: By education level | Overall | BMI (kg/m^2) | 28.85003 | 28.9 | mean1 |
| Panel B: By education level | < High school | BMI (kg/m^2) | 30.13442 | 30.1 | mean1 |
| Panel B: By education level | High school / GED | BMI (kg/m^2) | 30.24992 | 30.2 | mean1 |
| Panel B: By education level | Some college / AA | BMI (kg/m^2) | 30.62738 | 30.6 | mean1 |
| Panel B: By education level | College or higher | BMI (kg/m^2) | 28.27875 | 28.3 | mean1 |
| Panel B: By education level | Missing | BMI (kg/m^2) | 28.7563 | 28.8 | mean1 |
| Panel B: By education level | Overall | Current smoker (%) | 45.2652 | 45.30% | percent1 |
| Panel B: By education level | < High school | Current smoker (%) | 47.336 | 47.30% | percent1 |

| | | | | | |
|---|---|---|---|---|---|
| Panel B: By education level | High school / GED | Current smoker (%) | 47.15042 | 47.20% | percent1 |
| Panel B: By education level | Some college / AA | Current smoker (%) | 43.30242 | 43.30% | percent1 |
| Panel B: By education level | College or higher | Current smoker (%) | 22.90776 | 22.90% | percent1 |
| Panel B: By education level | Missing | Current smoker (%) | 46.10884 | 46.10% | percent1 |
| Panel B: By education level | Overall | Sugar-sweetened drink frequency (times/day) | 2.933075 | 2.9 | mean1 |
| Panel B: By education level | < High school | Sugar-sweetened drink frequency (times/day) | | | mean1 |
| Panel B: By education level | High school / GED | Sugar-sweetened drink frequency (times/day) | | | mean1 |
| Panel B: By education level | Some college / AA | Sugar-sweetened drink frequency (times/day) | | | mean1 |
| Panel B: By education level | College or higher | Sugar-sweetened drink frequency (times/day) | | | mean1 |
| Panel B: By education level | Missing | Sugar-sweetened drink frequency (times/day) | 2.933075 | 2.9 | mean1 |
| Panel B: By education level | Overall | Total sugar intake (g/day) | 113.7138 | 113.7 | mean1 |
| Panel B: By education level | < High school | Total sugar intake (g/day) | 94.71022 | 94.7 | mean1 |
| Panel B: By education level | High school / GED | Total sugar intake (g/day) | 101.3112 | 101.3 | mean1 |
| Panel B: By education level | Some college / AA | Total sugar intake (g/day) | 97.96532 | 98 | mean1 |
| Panel B: By education level | College or higher | Total sugar intake (g/day) | 90.1245 | 90.1 | mean1 |
| Panel B: By education level | Missing | Total sugar intake (g/day) | 115.7478 | 115.7 | mean1 |
| Panel B: By education level | Overall | Self-rated good oral health (%) | 72.24626 | 72.20% | percent1 |
| Panel B: By education level | < High school | Self-rated good oral health (%) | 49.39703 | 49.40% | percent1 |
| Panel B: By education level | High school / | Self-rated good oral health (%) | 62.52817 | 62.50% | percent1 |

| | | | | | |
|---|---|---|---|---|---|
| | **GED** | | | | |
| **Panel B: By education level** | **Some college / AA** | **Self-rated good oral health (%)** | **71.49051** | **71.50%** | **percent1** |
| **Panel B: By education level** | **College or higher** | **Self-rated good oral health (%)** | **88.44547** | **88.40%** | **percent1** |
| **Panel B: By education level** | **Missing** | **Self-rated good oral health (%)** | **72.1538** | **72.20%** | **percent1** |
| **Panel B: By education level** | **Overall** | **Past-year dental visit (%)** | **59.45722** | **59.50%** | **percent1** |
| **Panel B: By education level** | **< High school** | **Past-year dental visit (%)** | | | **percent1** |
| **Panel B: By education level** | **High school / GED** | **Past-year dental visit (%)** | | | **percent1** |
| **Panel B: By education level** | **Some college / AA** | **Past-year dental visit (%)** | | | **percent1** |
| **Panel B: By education level** | **College or higher** | **Past-year dental visit (%)** | | | **percent1** |
| **Panel B: By education level** | **Missing** | **Past-year dental visit (%)** | **59.45722** | **59.50%** | **percent1** |
| **Panel B: By education level** | **Overall** | **Unmet dental need (%)** | **18.7667** | **18.80%** | **percent1** |
| **Panel B: By education level** | **< High school** | **Unmet dental need (%)** | | | **percent1** |
| **Panel B: By education level** | **High school / GED** | **Unmet dental need (%)** | | | **percent1** |
| **Panel B: By education level** | **Some college / AA** | **Unmet dental need (%)** | | | **percent1** |
| **Panel B: By education level** | **College or higher** | **Unmet dental need (%)** | | | **percent1** |
| **Panel B: By education level** | **Missing** | **Unmet dental need (%)** | **18.7667** | **18.80%** | **percent1** |

“Additional sample characteristics by income and education are provided in Supplementary Table S1.”

---

**Table S2. Full regression models for DMFT and self-rated good oral health (NHANES)**

| Panel | Age gr | Predictor | Term | Estimat | Std_Erro | p_value | CI_low | CI_high | Odds_ratio | OR_CI_low | OR_CI_high |
|---|---|---|---|---|---|---|---|---|---|---|---|

| | | | | | | | | |
|---|---|---|---|---|---|---|---|---|
| | oup | | | e | r | | | |
| Panel A: DMFT | Overall | Intercept | Intercept | -4.83689 | 0.143971 | ###### | -5.11908 | -4.55471 |
| Panel A: DMFT | Overall | Education: College or higher vs < High school | C(education_std)[T.College+] | 1.741942 | 0.116541 | 1.63E-50 | 1.513522 | 1.970362 |
| Panel A: DMFT | Overall | Education: High school / GED vs < High school | C(education_std)[T.High school/GED] | 0.706673 | 0.122782 | 8.61E-09 | 0.466077 | 0.947382 |
| Panel A: DMFT | Overall | Education: Missing vs < High school | C(education_std)[T.Missing] | 4.832436 | 0.108688 | 0 | 4.619408 | 5.045464 |
| Panel A: DMFT | Overall | Education: Some college / AA vs < High school | C(education_std)[T.Some college/AA] | 1.161227 | 0.117355 | 4.38E-23 | 0.93121 | 1.391243 |
| Panel A: DMFT | Overall | Sex: Male vs Female | C(sex_label)[T.Male] | 0.02998 | 0.055451 | 0.588743 | -0.0787 | 0.138663 |
| Panel A: DMFT | Overall | Race/ethnicity: Missing vs reference | C(race_std)[T.Missing] | 1.827494 | 0.112072 | 8.89E-60 | 1.607832 | 2.047156 |
| Panel A: DMFT | Overall | Race/ethnicity: Non-Hispanic Black vs reference | C(race_std)[T.Non-Hispanic Black] | 1.164509 | 0.069435 | 3.96E-63 | 1.028417 | 1.300601 |
| Panel A: DMFT | Overall | Race/ethnicity: Non-Hispanic White vs reference | C(race_std)[T.Non-Hispanic White] | 0.643043 | 0.063979 | 9.11E-24 | 0.517644 | 0.768443 |

| | | | | | | | | |
|---|---|---|---|---|---|---|---|---|
| Panel A: DMFT | Overall | Race/ethnicity: Other Hispanic vs reference | C(race_std)[T.Other Hispanic] | 1.264153 | 0.094558 | 9.56E-41 | 1.078776 | 1.449529 |
| Panel A: DMFT | Overall | Race/ethnicity: Other/Multiracial vs reference | C(race_std)[T.Other/Multiracial] | -0.449322 | 0.152555 | 0.003226 | -0.748333 | -0.150331 |
| Panel A: DMFT | Overall | PIR (per 1-unit increase) | pir | -0.602055 | 0.018733 | ###### | -0.638877 | -0.565333 |
| Panel A: DMFT | Overall | Age (years) | age | 0.110744 | 0.001723 | 0 | 0.107366 | 0.114121 |
| Panel A: DMFT | Overall | Toothbrushing frequency (per day) | brush_freq_num | ###### | 3.13E-16 | 4.08E-10 | ###### | ###### |
| Panel A: DMFT | Overall | Coffee intake (mg/day) | coffee_mg_day | 0.001345 | 0.000192 | 2.38E-12 | 0.000969 | 0.001721 |
| Panel A: DMFT | Overall | Short sleep (<6h) | short_sleep | -1.055122 | 0.126542 | 7.55E-17 | -1.303314 | -0.8071 |
| Panel A: DMFT | Overall | Brush twice daily (=1) | brush_twice_daily | 0 | 0 | | 0 | 0 |
| Panel A: DMFT | Overall | Current smoker (=1) | smoke_current | 0 | 0 | | 0 | 0 |
| Panel A: DMFT | Ove | Caffeine 100 mg/day (=1) | caffeine_high_300mg | 0 | 0 | | 0 | 0 |

| | | | | | | | | |
|---|---|---|---|---|---|---|---|---|
| | rall | | | | | | | |
| Panel A: DMFT | Overall | Tea consumption (=1) | tea_any | 0 | 0 | | 0 | 0 |
| Panel A: DMFT | 20-44 | Intercept | Intercept | -1.657728 | 0.150894 | 4.61E-28 | -1.953033 | -1.361533 |
| Panel A: DMFT | 20-44 | Education: College or higher vs < High school | C(education_std)[T.College+] | 0.430688 | 0.080775 | 9.72E-08 | 0.272361 | 0.589 |
| Panel A: DMFT | 20-44 | Education: High school / GED vs < High school | C(education_std)[T.High school/GED] | 0.331273 | 0.083798 | 7.71E-05 | 0.167033 | 0.495517 |
| Panel A: DMFT | 20-44 | Education: Missing vs < High school | C(education_std)[T.Missing] | 1.994322 | 0.079848 | ###### | 1.837822 | 2.150825 |
| Panel A: DMFT | 20-44 | Education: Some college / AA vs < High school | C(education_std)[T.Some college/AA] | 0.413009 | 0.079705 | 2.20E-07 | 0.256787 | 0.569233 |
| Panel A: DMFT | 20-44 | Sex: Male vs Female | C(sex_label)[T.Male] | -0.190071 | 0.050052 | 0.000139 | -0.288811 | -0.092611 |
| Panel A: DMFT | 20-44 | Race/ethnicity: Missing vs reference | C(race_std)[T.Missing] | 0.709629 | 0.084356 | 4.02E-17 | 0.54429 | 0.874967 |
| Panel A: DMFT | 20-44 | Race/ethnicity: Non-Hispanic Black vs reference | C(race_std)[T.Non-Hispanic Black] | 0.326813 | 0.061848 | 1.26E-07 | 0.205591 | 0.448035 |

| | | | | | | | | |
|---|---|---|---|---|---|---|---|---|
| Panel A: DMFT | 20-44 | Race/ethnicity: Non-Hispanic White vs reference | C(race_std)[T.Non-Hispanic White] | 0.498707 | 0.065095 | 1.84E-14 | 0.371122 | 0.626293 |
| Panel A: DMFT | 20-44 | Race/ethnicity: Other Hispanic vs reference | C(race_std)[T.Other Hispanic] | 0.690378 | 0.09039 | 2.21E-14 | 0.513213 | 0.867543 |
| Panel A: DMFT | 20-44 | Race/ethnicity: Other/Multiracial vs reference | C(race_std)[T.Other/Multiracial] | -0.318811 | 0.129765 | 0.014229 | -0.572245 | -0.063377 |
| Panel A: DMFT | 20-44 | PIR (per 1-unit increase) | pir | -0.254833 | 0.018649 | 1.66E-42 | -0.291388 | -0.218288 |
| Panel A: DMFT | 20-44 | Age (years) | age | 0.078021 | 0.0038 | 1.09E-93 | 0.070574 | 0.085469 |
| Panel A: DMFT | 20-44 | Toothbrushing frequency (per day) | brush_freq_num | ###### | 1.26E-16 | 0.416815 | ###### | 1.45E-16 |
| Panel A: DMFT | 20-44 | Coffee intake (mg/day) | coffee_mg_day | 0.000914 | 0.000215 | 2.10E-05 | 0.000493 | 0.001334 |
| Panel A: DMFT | 20-44 | Short sleep (<6h) | short_sleep | -0.284877 | 0.112378 | 0.011248 | -0.505513 | -0.064646 |
| Panel A: DMFT | 20-44 | Brush twice daily (=1) | brush_twice_daily | 0 | 0 | | 0 | 0 |
| Panel A: DMFT | 20- | Current smoker (=1) | smoke_current | 0 | 0 | | 0 | 0 |

| | | | | | | | | |
|---|---|---|---|---|---|---|---|---|
| | 44 | | | | | | | |
| Panel A: DMFT | 20-44 | Caffeine 100 mg/day (=1) | caffeine_high_300mg | 0 | 0 | | 0 | 0 |
| Panel A: DMFT | 20-44 | Tea consumption (=1) | tea_any | 0 | 0 | | 0 | 0 |
| Panel A: DMFT | 45-64 | Intercept | Intercept | -7.44022 | 0.579306 | 9.38E-38 | -8.57566 | -6.30478 |
| Panel A: DMFT | 45-64 | Education: College or higher vs < High school | C(education_std)[T.College+] | 1.897792 | 0.165043 | 1.34E-30 | 1.574307 | 2.221277 |
| Panel A: DMFT | 45-64 | Education: High school / GED vs < High school | C(education_std)[T.High school/GED] | 0.453149 | 0.155909 | 0.003655 | 0.147567 | 0.758732 |
| Panel A: DMFT | 45-64 | Education: Missing vs < High school | C(education_std)[T.Missing] | 5.409177 | 0.155949 | ###### | 5.103517 | 5.714837 |
| Panel A: DMFT | 45-64 | Education: Some college / AA vs < High school | C(education_std)[T.Some college/AA] | 0.991514 | 0.153884 | 1.17E-10 | 0.689901 | 1.293127 |
| Panel A: DMFT | 45-64 | Sex: Male vs Female | C(sex_label)[T.Male] | 0.081388 | 0.111839 | 0.466783 | -0.13782 | 0.300593 |
| Panel A: DMFT | 45-64 | Race/ethnicity: Missing vs reference | C(race_std)[T.Missing] | 2.057607 | 0.21669 | 2.19E-21 | 1.632893 | 2.48232 |

| | | | | | | | | |
|---|---|---|---|---|---|---|---|---|
| Panel A: DMFT | 45-64 | Race/ethnicity: Non-Hispanic Black vs reference | C(race_std)[T.Non-Hispanic Black] | 1.767201 | 0.145732 | 7.66E-34 | 1.481565 | 2.052836 |
| Panel A: DMFT | 45-64 | Race/ethnicity: Non-Hispanic White vs reference | C(race_std)[T.Non-Hispanic White] | 0.888931 | 0.140424 | 2.45E-10 | 0.613701 | 1.164162 |
| Panel A: DMFT | 45-64 | Race/ethnicity: Other Hispanic vs reference | C(race_std)[T.Other Hispanic] | 1.674807 | 0.202123 | 1.17E-16 | 1.278645 | 2.070968 |
| Panel A: DMFT | 45-64 | Race/ethnicity: Other/Multiracial vs reference | C(race_std)[T.Other/Multiracial] | -0.440755 | 0.327363 | 0.178184 | -1.082238 | 0.200881 |
| Panel A: DMFT | 45-64 | PIR (per 1-unit increase) | pir | -0.791888 | 0.038415 | 2.05E-94 | -0.867177 | -0.716659 |
| Panel A: DMFT | 45-64 | Age (years) | age | 0.152752 | 0.009905 | 1.18E-53 | 0.133337 | 0.172167 |
| Panel A: DMFT | 45-64 | Toothbrushing frequency (per day) | brush_freq_num | 4.21E-15 | 3.89E-16 | 2.54E-27 | 3.45E-15 | 4.97E-15 |
| Panel A: DMFT | 45-64 | Coffee intake (mg/day) | coffee_mg_day | 0.001645 | 0.00032 | 2.70E-07 | 0.001018 | 0.002272 |
| Panel A: DMFT | 45-64 | Short sleep (<6h) | short_sleep | -1.098824 | 0.26741 | 4.01E-05 | -1.622336 | -0.574411 |
| Panel A: DMFT | 45-64 | Brush twice daily (=1) | brush_twice_daily | 0 | 0 | | 0 | 0 |

| | | | | | | | | |
|---|---|---|---|---|---|---|---|---|
| Panel A: DMFT | 45-64 | Current smoker (=1) | smoke_current | 0 | 0 | | 0 | 0 |
| Panel A: DMFT | 45-64 | Caffeine 100 mg/day (=1) | caffeine_high_300mg | 0 | 0 | | 0 | 0 |
| Panel A: DMFT | 45-64 | Tea consumption (=1) | tea_any | 0 | 0 | | 0 | 0 |
| Panel A: DMFT | 65+ | Intercept | Intercept | -5.363371 | 1.178576 | 5.34E-06 | -7.673372 | -3.0537 |
| Panel A: DMFT | 65+ | Education: College or higher vs < High school | C(education_std)[T.College+] | 2.591223 | 0.19784 | 3.40E-39 | 2.203457 | 2.97899 |
| Panel A: DMFT | 65+ | Education: High school / GED vs < High school | C(education_std)[T.High school/GED] | 1.183656 | 0.181316 | 6.66E-11 | 0.828276 | 1.539035 |
| Panel A: DMFT | 65+ | Education: Missing vs < High school | C(education_std)[T.Missing] | 8.663826 | 0.181908 | 0 | 8.307286 | 9.020367 |
| Panel A: DMFT | 65+ | Education: Some college / AA vs < High school | C(education_std)[T.Some college/AA] | 1.536952 | 0.184239 | 7.29E-17 | 1.175843 | 1.898061 |
| Panel A: DMFT | 65+ | Sex: Male vs Female | C(sex_label)[T.Male] | 0.424427 | 0.171754 | 0.013503 | 0.087632 | 0.760907 |
| Panel A: DMFT | 65+ | Race/ethnicity: Missing vs reference | C(race_std)[T.Missing] | 4.070482 | 0.464634 | 1.94E-18 | 3.1598 | 4.981163 |
| Panel A: | 6 | Race/ethnicity | C(race_std | 2. | 0. | 1. | 2. | 3. |

| | | | | | | | | |
|---|---|---|---|---|---|---|---|---|
| DMFT | 5+ | : Non-Hispanic Black vs reference | )[T.Non-Hispanic Black] | 781323 | 31609 | 38E-18 | 161786 | 400886 |
| Panel A: DMFT | 65+ | Race/ethnicity: Non-Hispanic White vs reference | C(race_std)[T.Non-Hispanic White] | 0.393474 | 0.278085 | 0.157085 | -0.151557 | 0.938521 |
| Panel A: DMFT | 65+ | Race/ethnicity: Other Hispanic vs reference | C(race_std)[T.Other Hispanic] | 2.443401 | 0.440585 | 2.93E-08 | 1.579855 | 3.306947 |
| Panel A: DMFT | 65+ | Race/ethnicity: Other/Multiracial vs reference | C(race_std)[T.Other/Multiracial] | -1.37704 | 0.758655 | 0.069507 | -2.86401 | 0.10992 |
| Panel A: DMFT | 65+ | PIR (per 1-unit increase) | pir | -1.06953 | 0.061 | 7.99E-69 | -1.18909 | -0.94997 |
| Panel A: DMFT | 65+ | Age (years) | age | 0.086946 | 0.015467 | 1.89E-08 | 0.056631 | 0.117262 |
| Panel A: DMFT | 65+ | Toothbrushing frequency (per day) | brush_freq_num | ###### | 2.06E-15 | 8.01E-12 | ###### | ###### |
| Panel A: DMFT | 65+ | Coffee intake (mg/day) | coffee_mg_day | 0.002273 | 0.000559 | 4.74E-05 | 0.001178 | 0.003369 |
| Panel A: DMFT | 65+ | Short sleep (<6h) | short_sleep | -3.32173 | 0.338706 | 1.05E-22 | -3.98559 | -2.65787 |
| Panel A: DMFT | 65+ | Brush twice daily (=1) | brush_twice_daily | 0 | 0 | | 0 | 0 |

| | | | | | | | | | | | |
|---|---|---|---|---|---|---|---|---|---|---|---|
| Panel A: DMFT | 65+ | Current smoker (=1) | smoke_current | 0 | 0 | | 0 | 0 | | | |
| Panel A: DMFT | 65+ | Caffeine 100 mg/day (=1) | caffeine_high_300mg | 0 | 0 | | 0 | 0 | | | |
| Panel A: DMFT | 65+ | Tea consumption (=1) | tea_any | 0 | 0 | | 0 | 0 | | | |
| Panel B: Self-rated good oral health (OR) | Overall | Intercept | Intercept | -0.535355 | 0.000493 | 0 | -0.536631 | -0.534438 | 0.585466 | 0.5849 | 0.586031 |
| Panel B: Self-rated good oral health (OR) | Overall | Education: College or higher vs < High school | C(education_std)[T.College+] | 1.238764 | 0.000549 | 0 | 1.237688 | 1.239839 | 3.451344 | 3.447635 | 3.455058 |
| Panel B: Self-rated good oral health (OR) | Overall | Education: High school / GED vs < High school | C(education_std)[T.High school/GED] | 0.241276 | 0.000494 | 0 | 0.240309 | 0.242244 | 1.272873 | 1.271641 | 1.274105 |
| Panel B: Self-rated good oral health (OR) | Overall | Education: Missing vs < High school | C(education_std)[T.Missing] | 0.551902 | 0.000426 | 0 | 0.551067 | 0.552736 | 1.736552 | 1.735104 | 1.738002 |
| Panel B: Self-rated good oral health (OR) | Overall | Education: Some college / AA vs < High school | C(education_std)[T.Some college/AA] | 0.461331 | 0.000498 | 0 | 0.460355 | 0.462306 | 1.586183 | 1.584637 | 1.587731 |
| Panel B: Self-rated good oral health (OR) | Overall | Sex: Male vs Female | C(sex_label)[T.Male] | -0.230085 | 0.00013 | 0 | -0.2311 | -0.230059 | 0.79386 | 0.793658 | 0.794061 |
| Panel B: Self-rated good oral health (OR) | Overall | Race/ethnicity: Missing vs reference | C(race_std)[T.Missing] | 0.549143 | 0.000299 | 0 | 0.548557 | 0.54973 | 1.731769 | 1.730754 | 1.732784 |
| Panel B: Self-rated good oral | Overall | Race/ethnicity: Non-Hispanic Black | C(race_std)[T.Non-Hispanic | 0.3615 | 0.0002 | 0 | 0.3610 | 0.3620 | 1.43556 | 1.43481 | 1.43631 |

| | | | | | | | | | | | |
|---|---|---|---|---|---|---|---|---|---|---|---|
| health (OR) | ll | vs reference | Black] | 6 | 67 | | 36 | 84 | 7 | 5 | 9 |
| Panel B: Self-rated good oral health (OR) | Overall | Race/ethnicity: Non-Hispanic White vs reference | C(race_std)[T.Non-Hispanic White] | 0.666511 | 0.000223 | 0 | 0.666073 | 0.666948 | 1.947443 | 1.946579 | 1.948282 |
| Panel B: Self-rated good oral health (OR) | Overall | Race/ethnicity: Other Hispanic vs reference | C(race_std)[T.Other Hispanic] | 0.314441 | 0.000302 | 0 | 0.313818 | 0.315002 | 1.369451 | 1.368864 | 1.370261 |
| Panel B: Self-rated good oral health (OR) | Overall | Race/ethnicity: Other/Multiracial vs reference | C(race_std)[T.Other/Multiracial] | 0.625922 | 0.000742 | 0 | 0.624468 | 0.627375 | 1.869968 | 1.867252 | 1.872689 |
| Panel B: Self-rated good oral health (OR) | Overall | PIR (per 1-unit increase) | pir | 0.351577 | 4.45E-05 | 0 | 0.35149 | 0.351664 | 1.421307 | 1.421183 | 1.421431 |
| Panel B: Self-rated good oral health (OR) | Overall | Age (years) | age | -0.005547 | 3.84E-06 | 0 | -0.005548 | -0.005546 | 0.994547 | 0.99454 | 0.994555 |
| Panel B: Self-rated good oral health (OR) | Overall | Toothbrushing frequency (per day) | brush_freq_num | ###### | 3.05E-21 | 0 | ###### | ###### | 1 | 1 | 1 |
| Panel B: Self-rated good oral health (OR) | Overall | Coffee intake (mg/day) | coffee_mg_day | -0.00071 | 3.18E-07 | 0 | -0.00071 | -0.00071 | 0.999293 | 0.999293 | 0.999294 |
| Panel B: Self-rated good oral health (OR) | Overall | Short sleep (<6h) | short_sleep | -0.536657 | 0.000247 | 0 | -0.53706 | -0.53609 | 0.58475 | 0.584466 | 0.585033 |
| Panel B: Self-rated good oral health (OR) | Overall | Brush twice daily (=1) | brush_twice_daily | 0 | 0 | | 0 | 0 | 1 | 1 | 1 |

| | | | | | | | | | | |
|---|---|---|---|---|---|---|---|---|---|---|
| Panel B: Self-rated good oral health (OR) | Overall | Current smoker (=1) | smoke_current | 0 | 0 | | 0 | 0 | 1 | 1 | 1 |
| Panel B: Self-rated good oral health (OR) | Overall | Caffeine 100 mg/day (=1) | caffeine_high_300mg | 0 | 0 | | 0 | 0 | 1 | 1 | 1 |
| Panel B: Self-rated good oral health (OR) | Overall | Tea consumption (=1) | tea_any | 0 | 0 | | 0 | 0 | 1 | 1 | 1 |
| Panel B: Self-rated good oral health (OR) | 20-44 | Intercept | Intercept | 0.284355 | 0.000873 | 0 | 0.282644 | 0.286067 | 1.328905 | 1.326632 | 1.331181 |
| Panel B: Self-rated good oral health (OR) | 20-44 | Education: College or higher vs < High school | C(education_std)[T.College+] | 1.548962 | 0.000906 | 0 | 1.547187 | 1.550737 | 4.706581 | 4.698233 | 4.714944 |
| Panel B: Self-rated good oral health (OR) | 20-44 | Education: High school / GED vs < High school | C(education_std)[T.High school/GED] | 0.282844 | 0.000813 | 0 | 0.281249 | 0.284438 | 1.326898 | 1.324784 | 1.329015 |
| Panel B: Self-rated good oral health (OR) | 20-44 | Education: Missing vs < High school | C(education_std)[T.Missing] | 0.53159 | 0.000712 | 0 | 0.530193 | 0.532986 | 1.701635 | 1.699261 | 1.704013 |
| Panel B: Self-rated good oral health (OR) | 20-44 | Education: Some college / AA vs < High school | C(education_std)[T.Some college/AA] | 0.583541 | 0.000818 | 0 | 0.581937 | 0.585144 | 1.792373 | 1.789501 | 1.79525 |
| Panel B: Self-rated good oral health (OR) | 20-44 | Sex: Male vs Female | C(sex_label)[T.Male] | -0.22932 | 0.000198 | 0 | -0.22971 | -0.22893 | 0.795075 | 0.794766 | 0.795384 |
| Panel B: Self-rated good oral health (OR) | 20-44 | Race/ethnicity: Missing vs reference | C(race_std)[T.Missing] | 0.602333 | 0.000422 | 0 | 0.601504 | 0.603157 | 1.826237 | 1.824861 | 1.82788 |
| Panel B: | 2 | Race/ethnicity | C(race_std | 0. | 0. | 0 | 0. | 0. | 1.5 | 1.5 | 1.5 |

| | | | | | | | | | | | |
|---|---|---|---|---|---|---|---|---|---|---|---|
| Self-rated good oral health (OR) | 0-44 | : Non-Hispanic Black vs reference | )[T.Non-Hispanic Black] | 462392 | 000372 | | 461663 | 46312 | 87867 | 86711 | 89024 |
| Panel B: Self-rated good oral health (OR) | 20-44 | Race/ethnicity: Non-Hispanic White vs reference | C(race_std)[T.Non-Hispanic White] | 0.482267 | 0.0003 | 0 | 0.481679 | 0.482856 | 1.619742 | 1.618789 | 1.620696 |
| Panel B: Self-rated good oral health (OR) | 20-44 | Race/ethnicity: Other Hispanic vs reference | C(race_std)[T.Other Hispanic] | 0.247361 | 0.000402 | 0 | 0.246574 | 0.248149 | 1.280642 | 1.279634 | 1.281651 |
| Panel B: Self-rated good oral health (OR) | 20-44 | Race/ethnicity: Other/Multiracial vs reference | C(race_std)[T.Other/Multiracial] | 1.208716 | 0.001317 | 0 | 1.206134 | 1.211298 | 3.349182 | 3.340546 | 3.357841 |
| Panel B: Self-rated good oral health (OR) | 20-44 | PIR (per 1-unit increase) | pir | 0.389263 | 7.13E-05 | 0 | 0.389123 | 0.389402 | 1.475892 | 1.475686 | 1.476098 |
| Panel B: Self-rated good oral health (OR) | 20-44 | Age (years) | age | -0.03099 | 1.40E-05 | 0 | -0.03102 | -0.03097 | 0.969481 | 0.969455 | 0.969508 |
| Panel B: Self-rated good oral health (OR) | 20-44 | Toothbrushing frequency (per day) | brush_freq_num | 2.66E-17 | 2.35E-20 | 0 | 2.66E-17 | 2.67E-17 | 1 | 1 | 1 |
| Panel B: Self-rated good oral health (OR) | 20-44 | Coffee intake (mg/day) | coffee_mg_day | -0.00043 | 5.45E-07 | 0 | -0.00043 | -0.00043 | 0.999572 | 0.999571 | 0.999573 |
| Panel B: Self-rated good oral health (OR) | 20-44 | Short sleep (<6h) | short_sleep | -0.50606 | 0.000379 | 0 | -0.5068 | -0.50531 | 0.602868 | 0.60242 | 0.603316 |
| Panel B: Self-rated good oral health (OR) | 20-44 | Brush twice daily (=1) | brush_twice_daily | 0 | 0 | | 0 | 0 | 1 | 1 | 1 |

| | | | | | | | | | | |
|---|---|---|---|---|---|---|---|---|---|---|
| Panel B: Self-rated good oral health (OR) | 20-44 | Current smoker (=1) | smoke_current | 0 | 0 | | 0 | 0 | 1 | 1 | 1 |
| Panel B: Self-rated good oral health (OR) | 20-44 | Caffeine 100 mg/day (=1) | caffeine_high_300mg | 0 | 0 | | 0 | 0 | 1 | 1 | 1 |
| Panel B: Self-rated good oral health (OR) | 20-44 | Tea consumption (=1) | tea_any | 0 | 0 | | 0 | 0 | 1 | 1 | 1 |
| Panel B: Self-rated good oral health (OR) | 45-64 | Intercept | Intercept | -0.4574 | 0.001266 | 0 | -0.45988 | -0.45492 | 0.632928 | 0.631359 | 0.6345 |
| Panel B: Self-rated good oral health (OR) | 45-64 | Education: College or higher vs < High school | C(education_std)[T.College+] | 1.128983 | 0.000933 | 0 | 1.127153 | 1.130812 | 3.092509 | 3.086857 | 3.098172 |
| Panel B: Self-rated good oral health (OR) | 45-64 | Education: High school / GED vs < High school | C(education_std)[T.High school/GED] | 0.064397 | 0.000822 | 0 | 0.062786 | 0.066008 | 1.066516 | 1.064799 | 1.068236 |
| Panel B: Self-rated good oral health (OR) | 45-64 | Education: Missing vs < High school | C(education_std)[T.Missing] | 0.456975 | 0.000703 | 0 | 0.455597 | 0.458353 | 1.579289 | 1.577115 | 1.581467 |
| Panel B: Self-rated good oral health (OR) | 45-64 | Education: Some college / AA vs < High school | C(education_std)[T.Some college/AA] | 0.405326 | 0.000826 | 0 | 0.403707 | 0.406946 | 1.499792 | 1.497365 | 1.502222 |
| Panel B: Self-rated good oral health (OR) | 45-64 | Sex: Male vs Female | C(sex_label)[T.Male] | -0.22509 | 0.000211 | 0 | -0.2255 | -0.22467 | 0.798446 | 0.798116 | 0.798777 |
| Panel B: Self-rated good oral health (OR) | 45-64 | Race/ethnicity: Missing vs reference | C(race_std)[T.Missing] | 0.444031 | 0.000521 | 0 | 0.443009 | 0.445052 | 1.558979 | 1.557387 | 1.560572 |

| | | | | | | | | | | | |
|---|---|---|---|---|---|---|---|---|---|---|---|
| Panel B: Self-rated good oral health (OR) | 45-64 | Race/ethnicity: Non-Hispanic Black vs reference | C(race_std)[T.Non-Hispanic Black] | 0.290077 | 0.000464 | 0 | 0.289168 | 0.290986 | 1.336531 | 1.335316 | 1.337746 |
| Panel B: Self-rated good oral health (OR) | 45-64 | Race/ethnicity: Non-Hispanic White vs reference | C(race_std)[T.Non-Hispanic White] | 0.657919 | 0.000396 | 0 | 0.657143 | 0.658696 | 1.930771 | 1.929273 | 1.93227 |
| Panel B: Self-rated good oral health (OR) | 45-64 | Race/ethnicity: Other Hispanic vs reference | C(race_std)[T.Other Hispanic] | 0.381315 | 0.000543 | 0 | 0.380251 | 0.382379 | 1.464209 | 1.462652 | 1.465768 |
| Panel B: Self-rated good oral health (OR) | 45-64 | Race/ethnicity: Other/Multiracial vs reference | C(race_std)[T.Other/Multiracial] | 0.417026 | 0.001078 | 0 | 0.414912 | 0.419139 | 1.517442 | 1.514238 | 1.520652 |
| Panel B: Self-rated good oral health (OR) | 45-64 | PIR (per 1-unit increase) | pir | 0.379824 | 6.91E-05 | 0 | 0.379688 | 0.379959 | 1.462027 | 1.461829 | 1.462225 |
| Panel B: Self-rated good oral health (OR) | 45-64 | Age (years) | age | -0.00957 | 1.87E-05 | 0 | -0.00961 | -0.00953 | 0.990476 | 0.99044 | 0.990512 |
| Panel B: Self-rated good oral health (OR) | 45-64 | Toothbrushing frequency (per day) | brush_freq_num | ###### | 1.51E-19 | 0 | ###### | ###### | 1 | 1 | 1 |
| Panel B: Self-rated good oral health (OR) | 45-64 | Coffee intake (mg/day) | coffee_mg_day | -0.0007 | 4.63E-07 | 0 | -0.0007 | -0.00069 | 0.999304 | 0.999304 | 0.999305 |
| Panel B: Self-rated good oral health (OR) | 45-64 | Short sleep (<6h) | short_sleep | -0.46292 | 0.00038 | 0 | -0.46367 | -0.46218 | 0.629441 | 0.628972 | 0.629911 |
| Panel B: | 4 | Brush twice | brush_twic | 0 | 0 | | 0 | 0 | 1 | 1 | 1 |

| | | | | | | | | | | | |
|---|---|---|---|---|---|---|---|---|---|---|---|
| Self-rated good oral health (OR) | 5-64 | daily (=1) | e_daily | | | | | | | | |
| Panel B: Self-rated good oral health (OR) | 45-64 | Current smoker (=1) | smoke_current | 0 | 0 | | 0 | 0 | 1 | 1 | 1 |
| Panel B: Self-rated good oral health (OR) | 45-64 | Caffeine 100 mg/day (=1) | caffeine_high_300mg | 0 | 0 | | 0 | 0 | 1 | 1 | 1 |
| Panel B: Self-rated good oral health (OR) | 45-64 | Tea consumption (=1) | tea_any | 0 | 0 | | 0 | 0 | 1 | 1 | 1 |
| Panel B: Self-rated good oral health (OR) | 65+ | Intercept | Intercept | -1.560037 | 0.002369 | 0 | -1.56502 | -1.555573 | 0.210058 | 0.209085 | 0.211035 |
| Panel B: Self-rated good oral health (OR) | 65+ | Education: College or higher vs < High school | C(education_std)[T.College+] | 0.697648 | 0.001057 | 0 | 0.695576 | 0.69972 | 2.009022 | 2.004864 | 2.013189 |
| Panel B: Self-rated good oral health (OR) | 65+ | Education: High school / GED vs < High school | C(education_std)[T.High school/GED] | 0.248625 | 0.000988 | 0 | 0.246688 | 0.250562 | 1.282261 | 1.27978 | 1.284747 |
| Panel B: Self-rated good oral health (OR) | 65+ | Education: Missing vs < High school | C(education_std)[T.Missing] | 0.624901 | 0.000836 | 0 | 0.623262 | 0.626541 | 1.868062 | 1.865002 | 1.871127 |
| Panel B: Self-rated good oral health (OR) | 65+ | Education: Some college / AA vs < High school | C(education_std)[T.Some college/AA] | 0.165991 | 0.000996 | 0 | 0.164037 | 0.167944 | 1.180562 | 1.178258 | 1.18287 |
| Panel B: Self-rated good oral health (OR) | 65+ | Sex: Male vs Female | C(sex_label)[T.Male] | -0.2399 | 0.000302 | 0 | -0.24049 | -0.23931 | 0.786706 | 0.786241 | 0.787172 |
| Panel B: | 6 | Race/ethnicity | C(race_std | 0. | 0. | 0 | 0. | 0. | 1.8 | 1.8 | 1.8 |

| | | | | | | | | | | | |
|---|---|---|---|---|---|---|---|---|---|---|---|
| Self-rated good oral health (OR) | 5+ | : Missing vs reference | )[T.Missing] | 619936 | 000853 | | 617688 | 621032 | 57739 | 546334 | 60848 |
| Panel B: Self-rated good oral health (OR) | 65+ | Race/ethnicity: Non-Hispanic Black vs reference | C(race_std)[T.Non-Hispanic Black] | 0.389445 | 0.000804 | 0 | 0.387869 | 0.391021 | 1.476162 | 1.473837 | 1.47849 |
| Panel B: Self-rated good oral health (OR) | 65+ | Race/ethnicity: Non-Hispanic White vs reference | C(race_std)[T.Non-Hispanic White] | 1.029465 | 0.000702 | 0 | 1.028089 | 1.03084 | 2.799567 | 2.795718 | 2.803421 |
| Panel B: Self-rated good oral health (OR) | 65+ | Race/ethnicity: Other Hispanic vs reference | C(race_std)[T.Other Hispanic] | 0.416182 | 0.000929 | 0 | 0.414361 | 0.418002 | 1.516161 | 1.513403 | 1.518924 |
| Panel B: Self-rated good oral health (OR) | 65+ | Race/ethnicity: Other/Multiracial vs reference | C(race_std)[T.Other/Multiracial] | 0.421465 | 0.001904 | 0 | 0.417732 | 0.425198 | 1.524193 | 1.518514 | 1.529893 |
| Panel B: Self-rated good oral health (OR) | 65+ | PIR (per 1-unit increase) | pir | 0.298678 | 0.000111 | 0 | 0.298461 | 0.298895 | 1.348076 | 1.347784 | 1.348369 |
| Panel B: Self-rated good oral health (OR) | 65+ | Age (years) | age | 0.009112 | 2.86E-05 | 0 | 0.009064 | 0.009176 | 1.009162 | 1.009105 | 1.009218 |
| Panel B: Self-rated good oral health (OR) | 65+ | Toothbrushing frequency (per day) | brush_freq_num | 3.92E-17 | 6.91E-20 | 0 | 3.91E-17 | 3.93E-17 | 1 | 1 | 1 |
| Panel B: Self-rated good oral health (OR) | 65+ | Coffee intake (mg/day) | coffee_mg_day | -0.00047 | 7.95E-07 | 0 | -0.00047 | -0.00046 | 0.999534 | 0.999533 | 0.999536 |
| Panel B: Self-rated good oral health (OR) | 65+ | Short sleep (<6h) | short_sleep | -0.50605 | 0.000671 | 0 | -0.50736 | -0.50473 | 0.602874 | 0.602081 | 0.603668 |
| Panel B: | 6 | Brush twice | brush_twic | 0 | 0 | | 0 | 0 | 1 | 1 | 1 |

| | | | | | | | | | | | |
|---|---|---|---|---|---|---|---|---|---|---|---|
| Self-rated good oral health (OR) | 5 + | daily (=1) | e_daily | | | | | | | | |
| Panel B: Self-rated good oral health (OR) | 65 + | Current smoker (=1) | smoke_current | 0 | 0 | | 0 | 0 | 1 | 1 | 1 |
| Panel B: Self-rated good oral health (OR) | 65 + | Caffeine 100 mg/day (=1) | caffeine_high_300mg | 0 | 0 | | 0 | 0 | 1 | 1 | 1 |
| Panel B: Self-rated good oral health (OR) | 65 + | Tea consumption (=1) | tea_any | 0 | 0 | | 0 | 0 | 1 | 1 | 1 |

**Table S3. Mediation analysis: indirect and direct effects of SES via dental access and sleep**

| Exposure | Exposure_ | Mediator | Mediator_ | Outcome | Outcome_ | Total_effe | Total_ci_lo | Total_ci_hi | Total_p_va | Total_effe | Total_or_c | Total_or_c | Direct_effe | Direct_ci_l | Direct_ci_h | Direct_p_v | Direct_effe | Direct_or_ | Direct_or_ | Indirect_ef | Indirect_ci | Indirect_ci | Indirect_p_ | Indirect_ef | Indirect_o | Indirect_o |
|---|---|---|---|---|---|---|---|---|---|---|---|---|---|---|---|---|---|---|---|---|---|---|---|---|---|---|
| PIR (per 1 | A=PIR | Past-year | visit_last_y | DMFT | dmft | -1.06798 | -1.12947 | -1.00648 | ###### | | | | -0.87539 | -0.93823 | -0.81255 | ###### | | | | -0.10387 | -0.11573 | -0.09484 | 0 | | | |
| PIR (per 1 | A=PIR | Past-year | visit_last_y | Self-rated | self_good | 0.395592 | 0.395484 | 0.395701 | 0 | 1.485264 | 1.485102 | 1.485425 | 0.329614 | 0.3295 | 0.329728 | 0 | 1.390431 | 1.390273 | 1.390589 | 0.0607 | 0.054966 | 0.065793 | 0 | 1.06258 | 1.056504 | 1.068005 |
| PIR (per 1 | A=PIR | Unmet de | unmet_ne | DMFT | dmft | -1.04907 | -1.11469 | -0.98344 | ###### | | | | -1.04042 | -1.10933 | -0.9715 | ###### | | | | -0.00255 | -0.01209 | 0.00744 | 0.608696 | | | |
| PIR (per 1 | A=PIR | Unmet de | unmet_ne | Self-rated | self_good | 0.395611 | 0.395501 | 0.39572 | 0 | 1.485291 | 1.485129 | 1.485453 | 0.298136 | 0.298022 | 0.298251 | 0 | 1.347346 | 1.347191 | 1.3475 | 0.089067 | 0.083812 | 0.095562 | 0 | 1.093154 | 1.087425 | 1.100277 |
| PIR (per 1 | A=PIR | Short slee | short_slee | DMFT | dmft | -1.0724 | -1.17536 | -0.96944 | 1.25E-92 | | | | -1.04768 | -1.15037 | -0.94498 | 6.04E-89 | | | | -0.01472 | -0.02275 | -0.0071 | 0.000226 | | | |
| PIR (per 1 | A=PIR | Short slee | short_slee | Self-rated | self_good | 0.359052 | 0.358923 | 0.35918 | 0 | 1.431971 | 1.431787 | 1.432154 | 0.348215 | 0.348086 | 0.348344 | 0 | 1.416537 | 1.416354 | 1.416719 | 0.010427 | 0.007808 | 0.013189 | 3.02E-14 | 1.010482 | 1.007839 | 1.013276 |

**Table S4. Oaxaca–Blinder decomposition: variable-level contributions to DMFT gap**

**内容：**

| Variable | Domain | Contribution | Share_of_gap | Share_of_gap_pct |
|---|---|---|---|---|
| visit_last_year | Access | -0.54918 | 0.417765 | 41.77648 |
| age_decade | Other | 0.544334 | -0.41408 | -41.4078 |
| unmet_need | Access | -0.24907 | 0.189473 | 18.94727 |
| education_group_Missing | Education | 0.152796 | -0.11623 | -11.6233 |
| race_std_Non-Hispanic Black | Race | -0.12278 | 0.093399 | 9.33994 |
| race_std_Non-Hispanic White | Race | 0.099424 | -0.07563 | -7.56328 |
| race_std_Other Hispanic | Race | -0.08501 | 0.064668 | 6.466837 |
| sleep_cat_Missing | Behavior | -0.03072 | 0.023366 | 2.336633 |
| sleep_cat_<6h | Behavior | -0.01719 | 0.013074 | 1.307372 |
| race_std_Missing | Race | -0.01362 | 0.010363 | 1.036324 |
| sex_label_Male | Other | -0.01055 | 0.008022 | 0.802175 |
| sleep_cat_>8h | Behavior | -0.00832 | 0.006331 | 0.633149 |
| education_group_Low | Education | 0.007093 | -0.0054 | -0.53956 |
| race_std_Other/Multiracial | Race | 0.0011 | -0.00084 | -0.08372 |
| education_group_High | Educatio | ###### | 7.03E-05 | 0.007025 |

| | n | | | |
|---|---|---|---|---|
| brush_freq_group_Missing | Behavior | 0 | 0 | 0 |
| brush_freq_group_>=3/day | Behavior | 0 | 0 | 0 |
| brush_freq_group_~2/day | Behavior | 0 | 0 | 0 |

**Table S5. Difference-in-differences sensitivity analyses for BRFSS outcomes**

Table S5. Difference-in-differences sensitivity checks for BRFSS senior outcomes (Dependent variables expressed in percentage points; clustered SE by state. All models include state and year fixed effects.)

| Outcome & Specification | β | SE | p-value |
|---|---|---|---|
| Dental visit rate (65+), baseline | -3.29 | 2.07 | 0.112 |
| Dental visit rate (65+), + state trends | -2.68 | 2.69 | 0.319 |
| Dental visit rate (65+), drop early expanders | -3.29 | 2.07 | 0.112 |
| Dental visit rate (65+), exclude partial expanders | -3.00 | 2.07 | 0.148 |
| Any tooth loss (65+), baseline | -1.06 | 1.73 | 0.539 |
| Any tooth loss (65+), + state trends | -0.09 | 2.33 | 0.968 |
| Any tooth loss (65+), drop early expanders | -1.06 | 1.73 | 0.539 |
| Any tooth loss (65+), exclude partial expanders | -1.02 | 1.76 | 0.563 |
| Complete edentulism (65+), baseline | -2.90 | 1.75 | 0.098 |
| Complete edentulism (65+), + state trends | 0.96 | 2.22 | 0.666 |
| Complete edentulism (65+), drop early expanders | -2.90 | 1.75 | 0.098 |
| Complete edentulism (65+), | -3.01 | 1.79 | 0.093 |

| |
|---|
| exclude partial expanders |

**Supplementary Figures**

**Supplementary Figure S1. Pearson correlation matrix for selected variables in NHANES adults.**

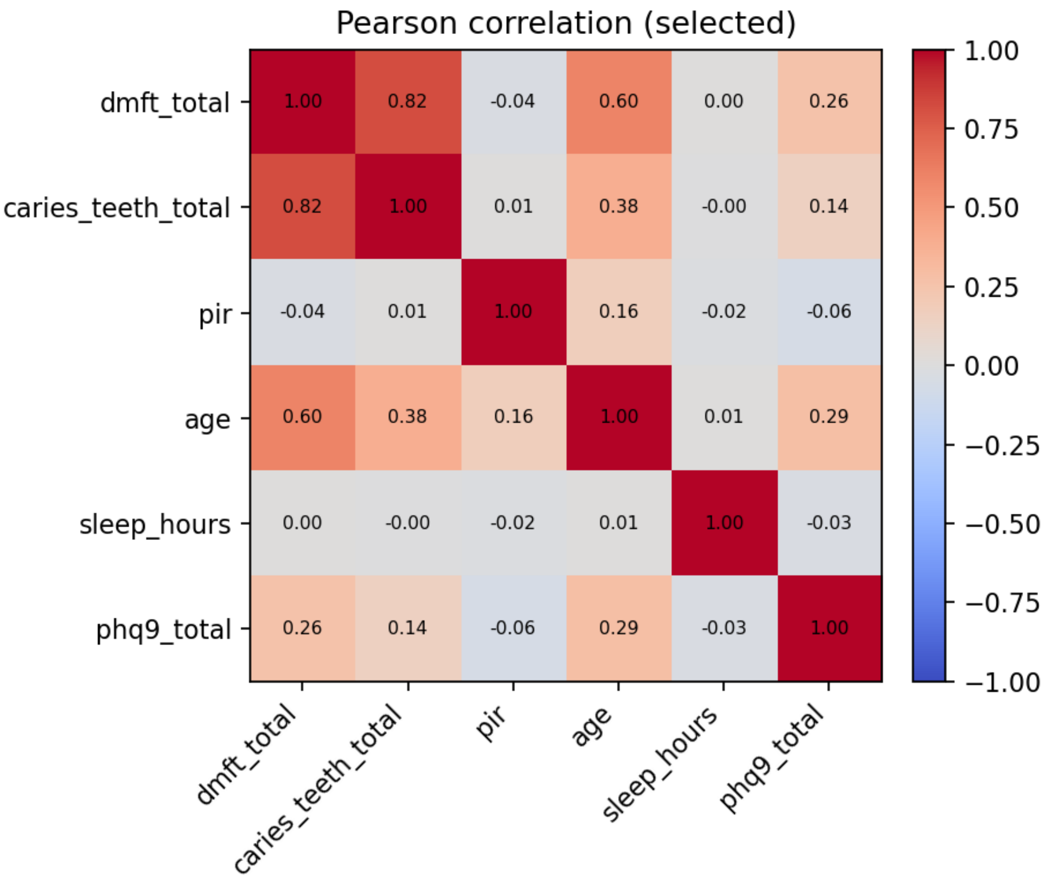

Notes: Heatmap of pairwise Pearson correlation coefficients among key variables used in the individual-level analyses: total DMFT (dmft_total), number of teeth with untreated caries (caries_teeth_total), poverty–income ratio (PIR), age, sleep duration in hours per night (sleep_hours) and depressive symptoms (PHQ-9 total score, phq9_total). Values in the cells indicate the correlation coefficient. Positive correlations are shown in red and negative correlations in blue, with stronger colours indicating stronger correlations. Estimates are based on survey-weighted NHANES data for adults aged ⩾20 years. **Supplementary Figure S2. Trends in dental visiting, unmet dental need and reasons by education and income, NHANES 1999–2020.**

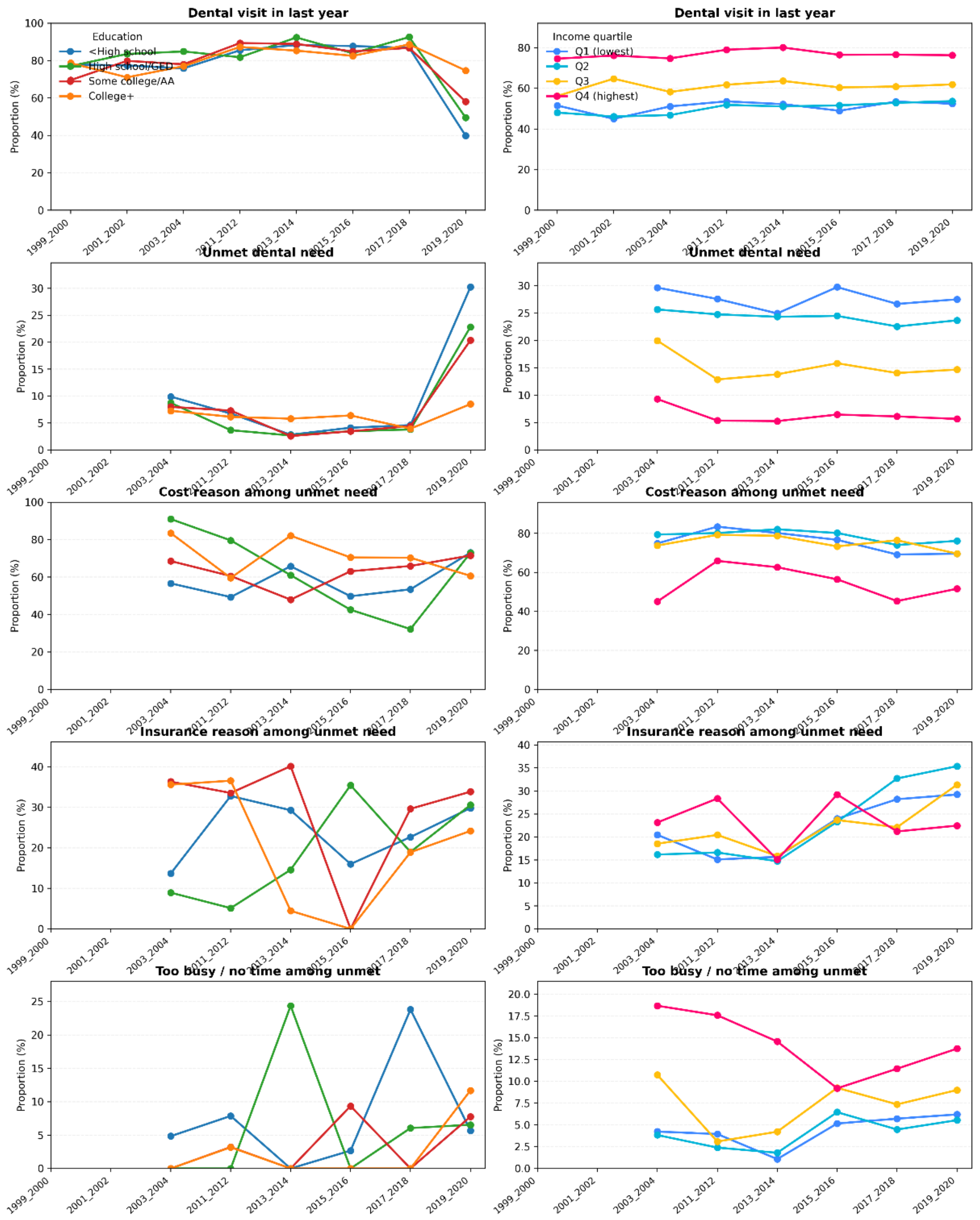

:20, NHANES 2003-2020. Proportions use interview weights. Education groups: <High school, High school/GED, Some college/AA, College+. Income groups: family PIR quartiles (Q1 lov

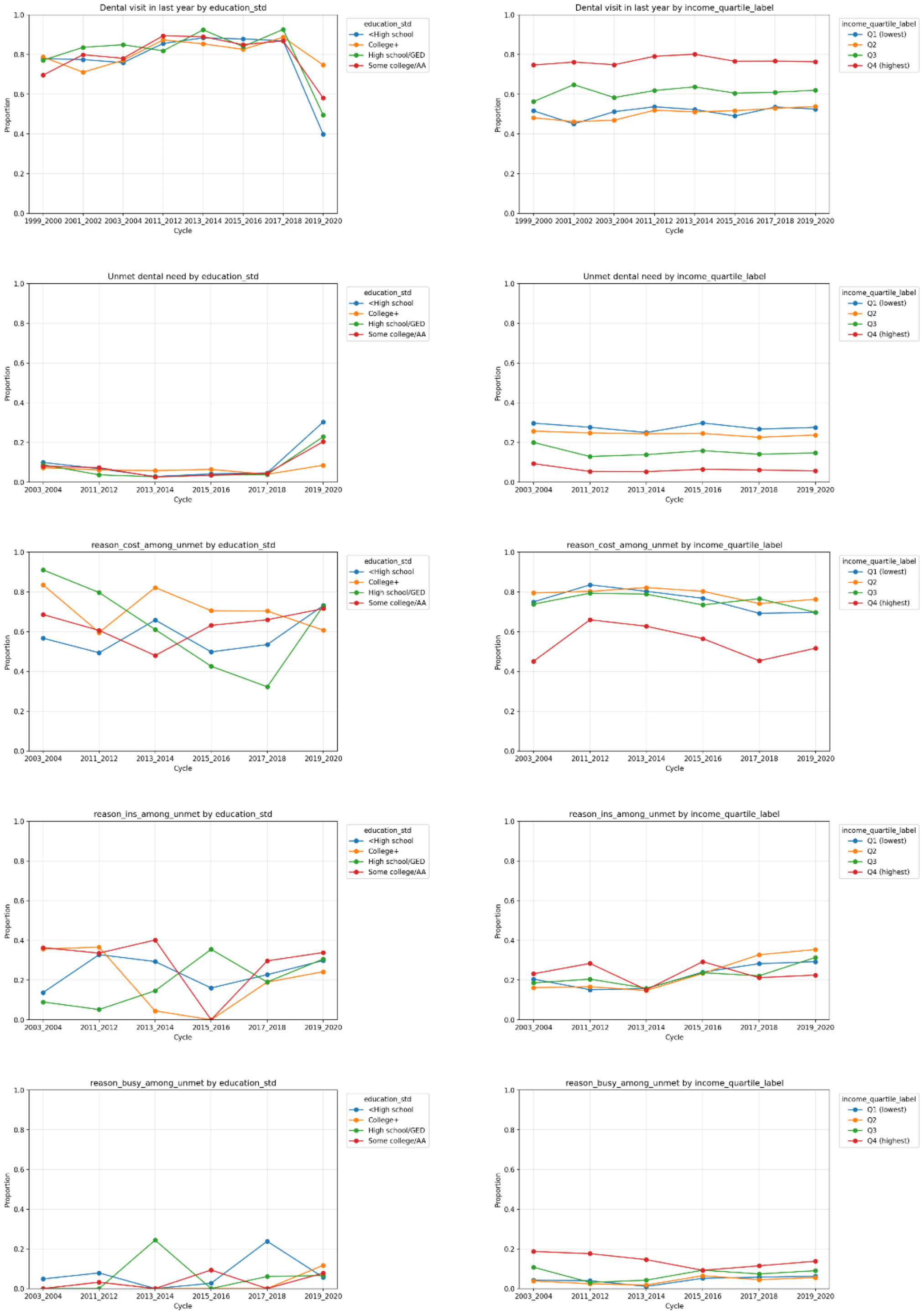

Notes: NHANES 2003-2020 adults age ≥20; estimates use interview weights.
Education: <High school, High school/GED, Some college/AA, College+.
Income: family PIR quartiles (Q1 lowest to Q4 highest).

**Notes:** Panels on the left show education groups (<High school, High school/GED, Some college/associate degree, College+); panels on the right show family poverty–income ratio (PIR) quartiles (Q1 lowest to Q4 highest). The first row presents the proportion of adults reporting a dental visit in the last year. The second row shows the proportion reporting any unmet dental need due to cost. The third row displays, among those with unmet need, the proportion citing cost as a reason. The fourth row displays, among those with unmet need,

the proportion citing lack of insurance or being too busy / having no time as a reason. Estimates are survey-weighted and calculated for adults aged ⩾20 years in each 2-year NHANES cycle (1999–2000 to 2019–2020).

**Supplementary Figure S3. Dental visiting, unmet dental need and reasons by education and income, NHANES 2003–2020.**

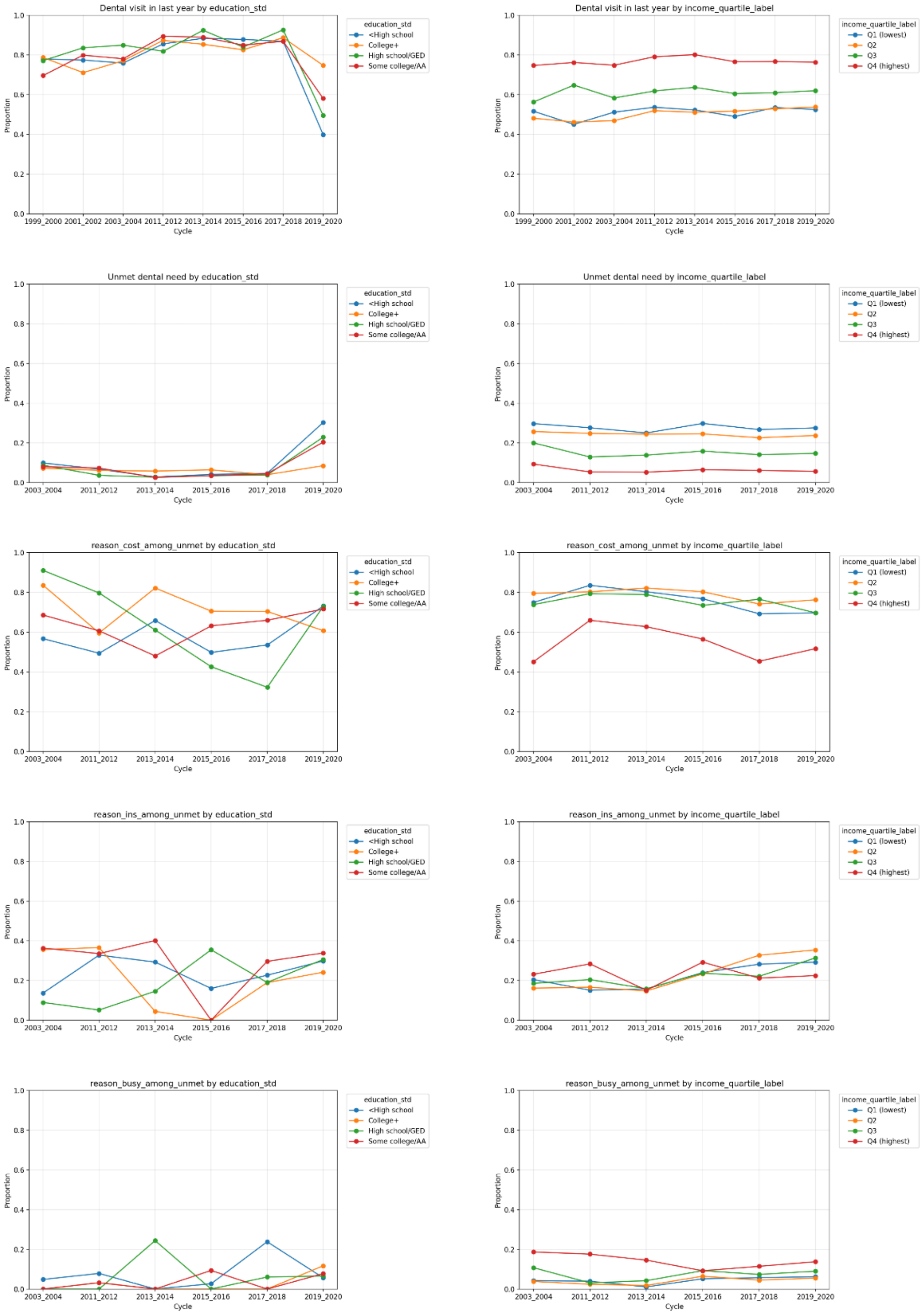

Notes: NHANES 2003-2020 adults age ≥20; estimates use interview weights.
Education: <High school, High school/GED, Some college/AA, College+.
Income: family PIR quartiles (Q1 lowest to Q4 highest).

Notes: Panels on the left show education groups (<High school, High school/GED, Some college/associate degree, College+); panels on the right show family poverty–income ratio (PIR) quartiles (Q1 lowest to Q4 highest). The first row presents the proportion of adults reporting a dental visit in the last year. The second row shows the proportion reporting any unmet dental need due to cost. The third row shows, among those with unmet need, the proportion citing cost as a reason. The fourth row shows, among those with unmet need, the proportion citing lack of insurance or being too busy / having no time as a reason. Estimates are based on NHANES 2003–2020 adults aged ≥20 years and use interview weights; proportions are calculated within each 2-year survey cycle.

**Supplementary Figure S4. Time trends in any dental caries by education, income and race/ethnicity, NHANES 1999–2020.**

**Notes:** Survey-weighted prevalence of any dental caries (%) among adults aged ≥20 years in NHANES 1999–2020, by 2-year survey cycle. (A) Prevalence by education level (<High school, High school/GED, Some college/associate degree, College+). (B) Prevalence by family poverty–income ratio (PIR) quartiles (Q1 lowest to Q4 highest). (C) Prevalence by race/ethnicity (Mexican American, non-Hispanic Black, non-Hispanic White, other Hispanic, other/multiracial). Lines connect point estimates across cycles to aid visualisation of trends; all estimates use examination weights and account for the complex survey design.

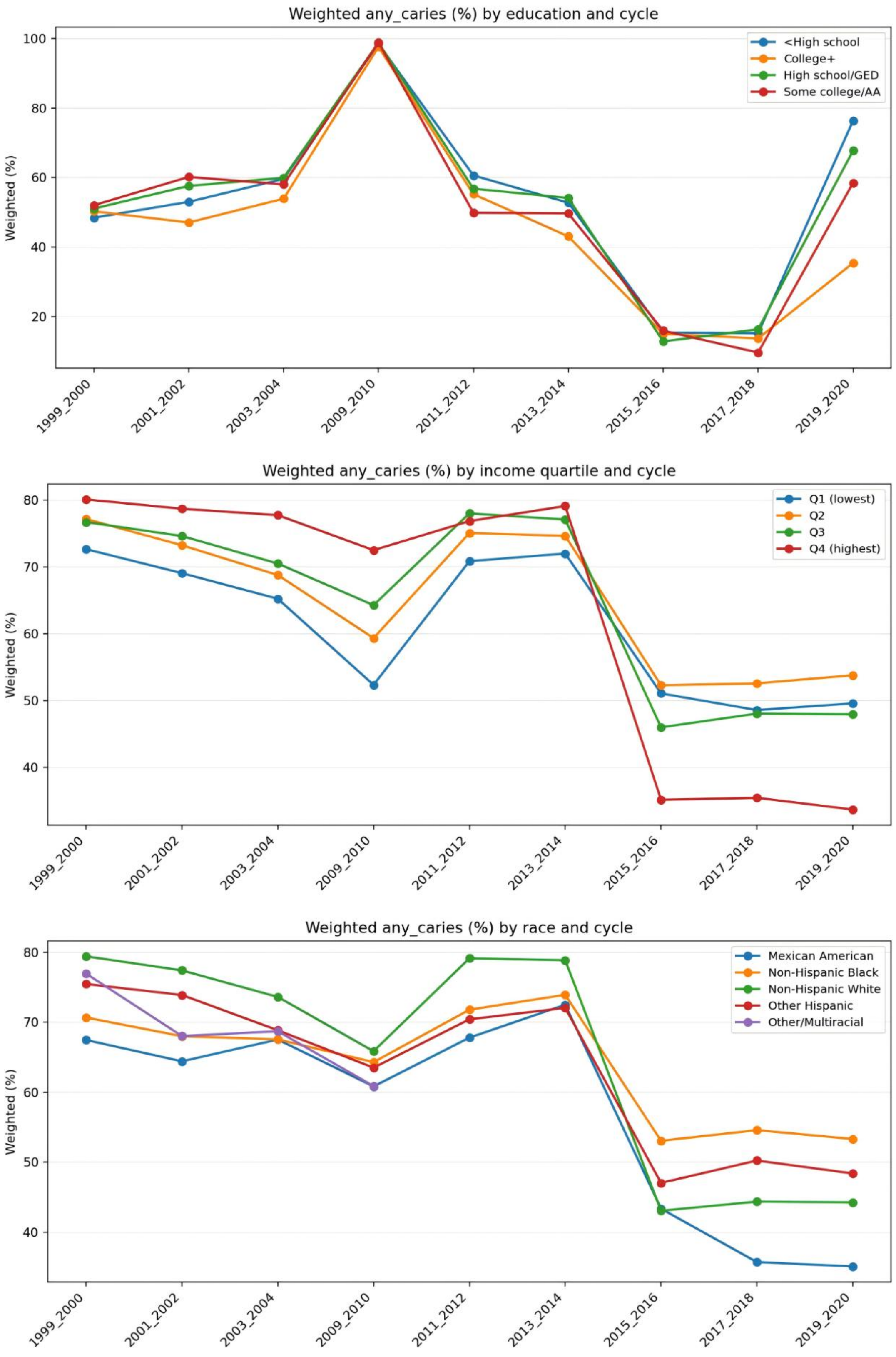

**Figure S5. BRFSS trends stratified by adult dental benefit generosity level**

Figure S5. Oral health trends by dental benefit generosity level

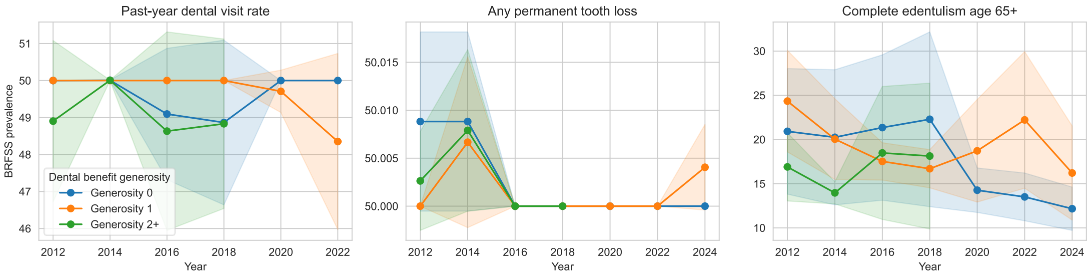
